# Ultra-rare coding variants in schizophrenia risk genes are associated with longitudinal phenotypic differences across psychotic disorders

**DOI:** 10.64898/2026.10.30.26366332

**Authors:** Anders Kämpe, Julia Sealock, Evan J. Giangrande, Calwing Liao, Emilia Vartiainen, Jaana Suvisaari, Markku Lähteenvuo, Tiina Paunio, Lea Urpa, Olli Kampman, Annamari Tuulio-Henriksson, Jouko Lönnqvist, Erkki Isometsä, Tuula Kieseppä, Kaisla Lahdensuo, Tuomo Kiiskinen, SUPERFinland-Researchers, Eija Hämäläinen, Mark J. Daly, Benjamin M. Neale, Aarno Palotie, Olli Pietiläinen

## Abstract

**Background:** The Schizophrenia Exome Meta-Analysis (SCHEMA) Consortium has recently identified 40 genes at a false discovery rate <0.05 in which ultra-rare damaging variants substantially increase schizophrenia (SZ) risk. However, the impact of these variants on clinical trajectories across the psychosis spectrum is not yet known.

**Methods:** We screened 9,821 sequenced participants with psychotic disorders who passed genomic quality control from the SUPER-Finland psychosis study for ultra-rare variants predicted to be damaging in these 40 SZ-associated genes, identifying 163 variant carriers. To assess the impact of these variants on clinical course and functioning across psychotic disorders, we used five decades of nationwide, full-coverage health register data together with interview, questionnaire, and cognitive assessments collected at study enrollment. We compared the clinical trajectories and functional outcomes of these variant carriers with those of non-carriers, including comparisons across different strata of the schizophrenia polygenic risk score (SZ-PGS) and with participants harboring deleterious variants in other genes associated with neurodevelopmental disorders (NDDs).

**Results:** Overall, the variant carriers had a similar distribution of major psychotic disorder diagnoses to that of non-carriers. Despite this diagnostic similarity, variant carriers had earlier hospital admissions than non-carriers for both psychiatric (HR=2.30 [1.48-3.57], p=2.05e-04) and non-psychiatric diagnoses (HR=1.40 [1.10-1.79], p=6.88e-03), indicating increased medical needs early in life. They also showed greater exposure to childhood adversity, lower attainment of adult social milestones, and poorer neuropsychological test performance at study inclusion. Across longitudinal measures, variant carriers showed less favorable clinical and functional trajectories than non-carriers and clustered more closely with individuals with high SZ-PGS (≥90th percentile). Consistent with a liability-threshold model, the variant carriers had lower SZ-PGS than non-carriers among participants with a diagnosis of SZ (β = -0.40, p = 5.32e-05) but higher SZ-PGS than population controls (β = 0.55, p = 9.36e-08), suggesting that the variant carriers developed psychosis at lower levels of common-variant liability. Gene-level analyses identified associations of variants in *AKAP11*, *SETD1A*, and *XPO7* with lower SZ-PGS and/or poorer neuropsychological test performance, while carriers of damaging variants in SZ-risk genes also associated with bipolar disorder showed high cumulative burdens of affective disorder diagnoses.

**Conclusion:** The disease trajectories of carriers of ultra-rare damaging variants in SZ-risk genes broadly overlapped with those of non-carriers, but carriers showed several differences: greater childhood adversity, earlier hospitalizations, and poorer neuropsychological test performance. These carrier-associated differences were detectable across psychotic disorder diagnoses and suggest a broader phenotypic impact of rare SZ-risk variants than is captured by SZ diagnosis alone, with potential clinical relevance for future risk stratification and genetic counseling.

## Introduction

Schizophrenia (SZ) is a psychotic disorder affecting multiple domains of behavior and cognitive functioning and is often regarded as a prototypical disorder within the psychosis spectrum (1). SZ is highly heritable (H^2^ ≈ 80%) (2) and has a complex genetic architecture likely involving thousands of common alleles with small effects (3) and ultra-rare variants with large effects on disease risk, including copy number variants (CNVs) (4) and deleterious coding variants (5, 6). Psychotic disorders vary considerably in their clinical courses, with SZ typically associated with the poorest outcomes (7). In addition to capturing disease liability, the common-variant polygenic score for SZ (SZ-PGS) has been associated with clinically relevant outcomes, including diagnostic shifts, need for hospital care, and global functioning (8, 9). However, individuals with the same overall SZ-PGS may carry different combinations of contributing alleles, and the aggregate measure itself offers only limited insight into the underlying biological mechanisms (3). Moreover, many of the clinical features observed in SZ are shared across psychotic disorders (8), making individual-level prediction of disease trajectories from early symptoms challenging.

In the recent 2026 update, the Schizophrenia Exome Meta-Analysis (SCHEMA) Consortium, to which the SUPER-Finland psychosis study contributed, analyzed 87,959 cases and 150,587 controls and identified 40 genes in which variants predicted to be damaging were associated with SZ at a false discovery rate <0.05 (10) [Methods]. These high-risk variants confer substantially increased risk for SZ and directly implicate specific genes in disease etiology, creating new opportunities for early identification of individual disease liability. Here we explored how these ultra-rare damaging variants influence disease course, symptom profiles, and comorbidities across the psychotic disorder spectrum.

We combined five decades of nationwide health register data with interview, questionnaire, and cognitive assessment data from 9,821 sequenced individuals with psychotic disorders who passed genomic quality control in the SUPER-Finland psychosis study (11). Because several SCHEMA-nominated genes have also been implicated in neurodevelopmental disorders (NDDs), we additionally compared SCHEMA variant carriers with participants carrying disruptive variants in NDD-associated genes not nominated by SCHEMA. Importantly, the broad recruitment of SUPER-Finland across psychotic disorders allowed us to assess the effects of damaging variants in SZ-risk genes beyond conventional diagnostic boundaries, which may otherwise obscure the full phenotypic spectrum of these variants and limit our understanding of their penetrance across different outcomes.

## Results

### Variant carriers had similar demographic and diagnostic profiles as non-carriers

We screened sequence data from 9,821 SUPER-Finland participants with a psychotic disorder who passed genomic quality control (QC) for variants predicted to be damaging in the 40 SCHEMA-nominated SZ-risk genes (median OR=4.53 for SZ) (10). In total, we identified 163 participants (1.7%) – hereafter called variant carriers – harboring variants predicted to be damaging in 38 of the 40 SCHEMA-nominated SZ-risk genes. Most variant carriers (n = 155) harbored predicted loss-of-function (pLoF) variants, and eight harbored predicted damaging missense variants (AlphaMissense score ≥0.98 and MPC score ≥3) [Methods]. Of the 163 variant carriers, 70 had damaging variants in 15 of the 16 genes that reached exome-wide significance for association with SZ (p<1.31e-06) in the SCHEMA analysis (10). The remaining 93 variant carriers had damaging variants in 23 of the 24 genes associated with SZ at a 5% false discovery rate (FDR < 0.05; p<6.84e-05) [Table S1a-b]. Overall, variant carriers and non-carriers (n=9,658 after QC) had similar demographic characteristics and distributions of psychotic disorder diagnoses [**Fig. 1a**], indicating that carrier status was not confined to a single diagnostic category. For carriers with damaging variants in one of the 15 exome-wide significant genes (n=70), a diagnosis of SZ was more frequent than among non-carriers in SUPER-Finland, although the difference was not significant (62.9% vs. 55.7%, p=0.22) [Table S2]. In comparison, 69.4% of non-carriers in the highest SZ-PGS decile had a diagnosis of SZ, whereas only 37.7% of those in the lowest SZ-PGS decile had a diagnosis of SZ [Fig. S1].

**Figure 1.**
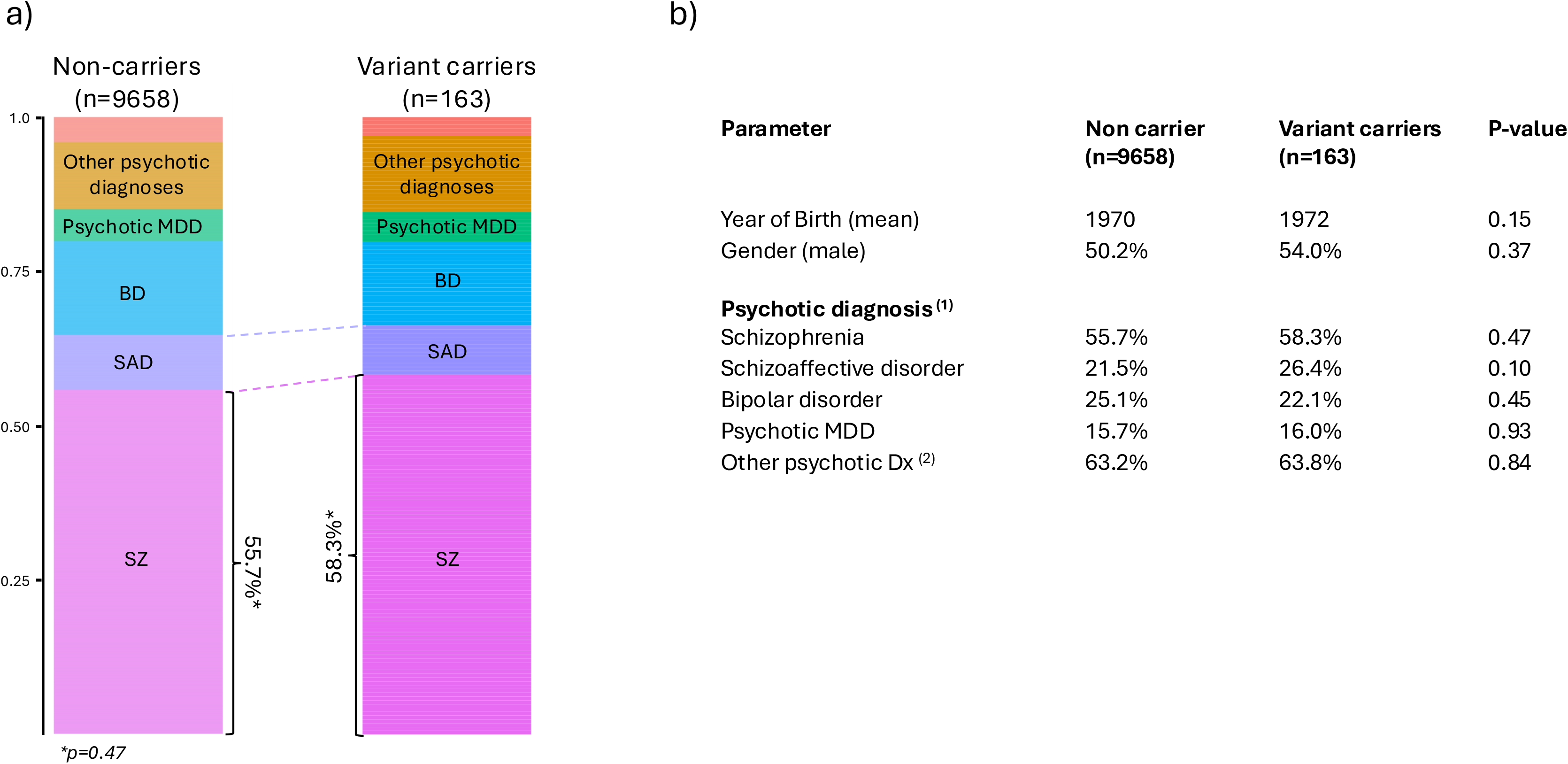
Diagnostic composition of variant carriers in SUPER-Finland. **a**) Distribution of major lifetime psychotic diagnoses among variant carriers and non-carriers in SUPER-Finland. The top segment represents participants without a register-recorded psychotic diagnosis. **b)** Variant carriers and non-carriers showed similar demographic characteristics and frequencies of psychotic diagnoses recorded at any time. *^(1)^ All psychotic diagnoses recorded at any time were considered; therefore, a single individual could contribute to multiple diagnostic categories.* ^(2)^ *‘Other psychotic diagnoses’ were broadly defined to include any diagnosis indicating a psychotic episode, although the majority consisted of diagnoses equivalent to F21-F24 and F26-F29*.

### Damaging variants in SZ-risk genes support a liability-threshold model for psychotic disorders

We first analyzed the polygenic background of the variant carriers. Consistent with a liability-threshold model, variant carriers with a diagnosis of SZ (n=95) had lower SZ-PGS than non-carriers with SZ (0.55 SD vs 0.95 SD, β= -0.40, p=5.32e-05), but higher SZ-PGS than population controls (β= 0.55, p= 9.36e-08) [**Fig. 2a**]. The association between variant carrier status and lower SZ-PGS also remained evident without stratification by SZ diagnosis (β= -0.32, p=2.47e-05). We also observed a negative association between the gene-wise SCHEMA-reported effect sizes and SZ-PGS among the variant carriers (β = -0.30, p=3.41e-03). To examine this relationship further, we ranked the variant carriers by the genes’ SCHEMA-reported effect sizes and divided them at the median into high-and low-effect-size groups. Of the variant carriers with a diagnosis of SZ, 51 were in the high-effect-size group (median OR = 7.30) and 44 were in the low-effect-size group (median OR = 2.68) [Table S3]. We then compared SZ-PGS in the high- and low-effect-size groups with that in non-carriers with a diagnosis of SZ. Variant carriers in the high-effect-size group showed a notably larger reduction in SZ-PGS relative to non-carriers (0.29 SD vs 0.95 SD, β= -0.63, p=2.88e-06) than those in the low-effect-size group (0.87 SD vs 0.95 SD, β= -0.14, p=0.35) [Fig. S2]. This dose-related pattern further supports the liability-threshold model, with larger SCHEMA-reported gene-level effect sizes corresponding to lower SZ-PGS among carriers with a diagnosis of SZ.

**Figure 2.**
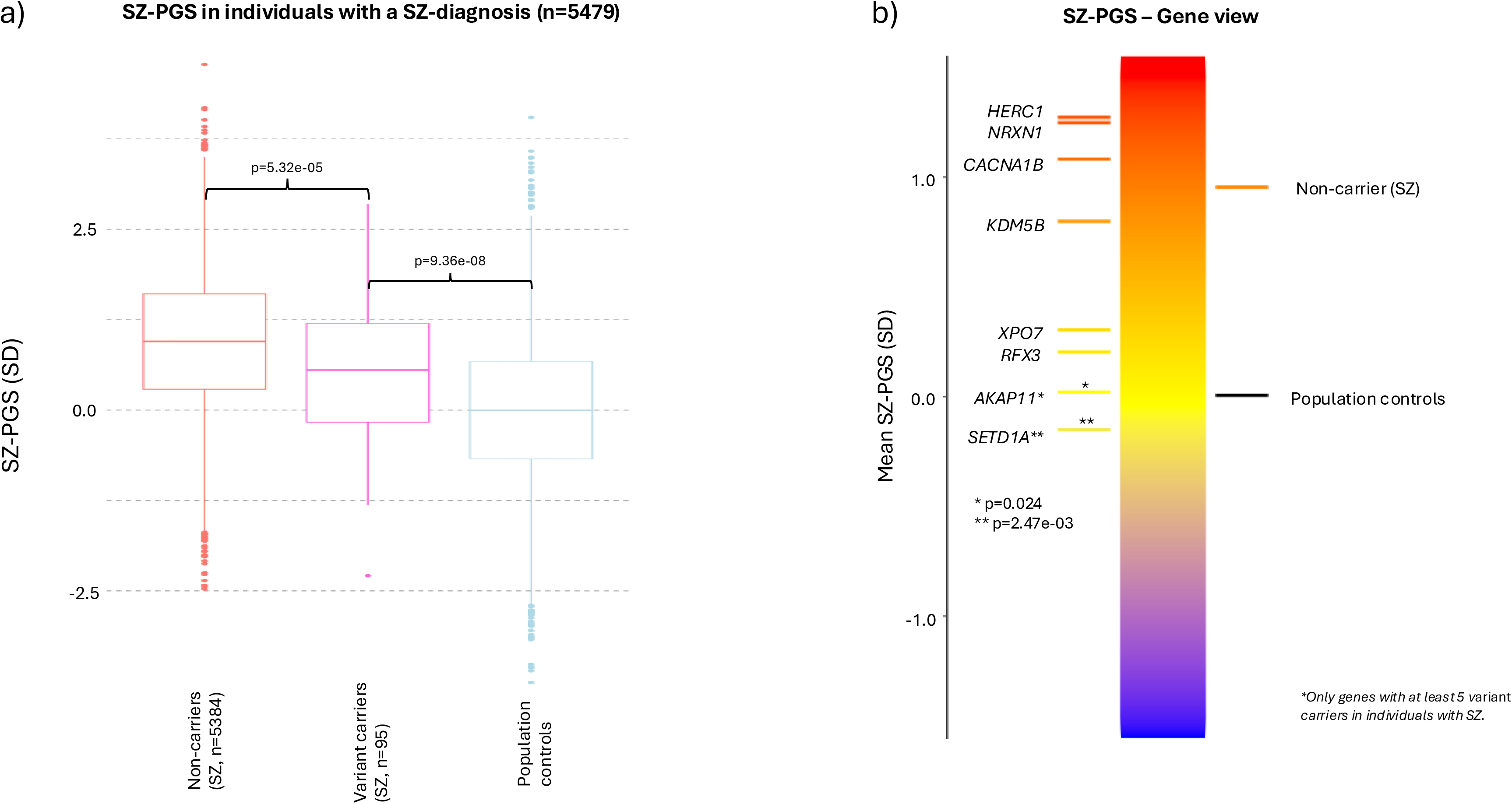
SZ-PGS in variant carriers supports a liability-threshold model. **a)** SZ-PGS among variant carriers compared with non-carriers and population controls. The analysis was restricted to individuals with a diagnosis of SZ. **b)** Mean SZ-PGS by SZ-risk gene among genes with at least five variant carriers with a diagnosis of SZ. *SETD1A* (p= 2.47e-03, FDR<0.05) and *AKAP11 (*p=0.024, nominally significant) were individually associated with a lower SZ-PGS.

Gene-level differences in polygenic background were also observed. Focusing on SCHEMA-nominated genes with at least 5 variant carriers among individuals with a diagnosis of SZ (n = 8 genes), we observed that *SETD1A* variant carriers (n=9 carriers) had a significantly lower SZ-PGS than non-carriers with a diagnosis of SZ (mean SZ-PGS: -0.16 SD vs 0.95 SD, p=2.47e-03, FDR<0.05), while *AKAP11* variant carriers had nominally lower SZ-PGS (p=0.024) [**Fig. 2b**]. The gene-level SZ-PGS results were further supported by empirical resampling analyses across 10,000 iterations (*SETD1A*, empirical p = 2.38e-03; *AKAP11*, empirical p = 0.022).

#### The liability-threshold model distinguishes variant carriers from non-carriers with NDD variants

The SCHEMA-nominated SZ-risk genes have a notable overlap with genes in which deleterious variants have previously been linked to neurodevelopmental disorders (NDDs) (10) [Details in Table S4]. This overlap prompted us to ask whether the reduced SZ-PGS observed in variant carriers with a SZ diagnosis was specific to damaging variants in SCHEMA-nominated SZ-risk genes or was also observed in individuals with disruptive variants in other NDD-associated genes. We therefore screened the sequence data for disruptive variants in haploinsufficient genes associated with dominant NDD syndromes but not implicated by SCHEMA, identifying 87 non-carriers with disruptive variants in NDD genes, including 54 with a SZ diagnosis [Methods]. These individuals are hereafter referred to as non-carriers with NDD variants. In contrast to variant carriers in SCHEMA-nominated genes, non-carriers with NDD variants and a diagnosis of SZ had a mean SZ-PGS as high as that of other non-carriers with the same diagnosis (0.92 SD vs 0.95 SD, p=0.99) [Fig. S3].

To further explore the relationship between neurodevelopmental disorders, psychosis, and SZ-PGS, we additionally leveraged data from the Northern Finnish Intellectual Disability Cohort (NFID), encompassing 1,139 index patients ascertained for ID (12). Compared with NFID participants, non-carriers with NDD variants in SUPER-Finland had higher SZ-PGS (β = 0.44 [0.21–0.67], p=1.72e-04) [Fig. S3], consistent with the different diagnostic ascertainment. Conversely, NFID participants who also had a diagnosis of a psychotic disorder (n=141, 12.4%) had a SZ-PGS similar to that of SUPER-Finland participants (β= -0.06 [-0.25–0.13], p= 0.53), while NFID participants without a reported psychotic episode diagnosis had markedly lower SZ-PGS than participants in SUPER-Finland (β= -0.55, p= 7.64e-61). Together, these results suggest that the reduced SZ-PGS observed in SCHEMA variant carriers is not a general feature of psychosis in individuals with NDD-associated variants or intellectual disability.

### Variant carriers showed greater early-life adversity

To understand whether variant carrier status was associated with differences before, around, or after psychotic illness onset, we leveraged Finland’s longitudinal medical registers, which include nationwide inpatient records since 1969, together with interview and questionnaire data collected at study enrollment. These data allowed us to assess when differences between variant carriers and non-carriers first emerged. This life-course analysis focused on three periods of life: 1) childhood and elementary school years (0–15 years); 2) early adulthood, covering attainment of adult social milestones and onset of psychotic illness; and 3) later adulthood, covering post-diagnosis and current everyday functioning, including cognitive assessments [Methods]. Childhood measures included both register-based and self-reported information on childhood family circumstances, childhood wellbeing, school performance, age at first non-psychiatric hospitalization, age at first psychiatric hospitalization, and hospital burden before the first recorded psychotic diagnosis [Supp. Appendix pp. 9–11].

The variant carriers had less favorable clinical and functional trajectories compared with non-carriers, and were most similar to individuals with the highest SZ-PGS (≥90^th^ percentile) [**Fig. 3**]. However, the variant carriers primarily showed less favorable outcomes during childhood and adolescence, before their first registry-recorded psychotic episode. Individuals with high SZ-PGS, on the other hand, displayed differences from other non-carriers primarily after psychiatric illness onset [Table S6]. In a direct comparison between variant carriers and the high SZ-PGS group, we used bootstrap resampling to test whether effect sizes differed between the two groups across the longitudinal measures [Supp. Appendix pp. 12]. This analysis indicated that the variant carriers showed larger differences than the high SZ-PGS group for self-reported childhood wellbeing, self-reported school performance, and age at first non-psychiatric hospitalization (FDR<0.05, Fig. S4). As a complementary analysis we repeatedly downsampled the high SZ-PGS group to match the sample size of the variant carriers (n=163). In this analysis of the high SZ-PGS group, only the association with age at first recorded psychotic diagnosis remained FDR-significant in at least 80% of resampling iterations (89.3%) [Supp. Appendix, pp. 13].

**Figure 3.**
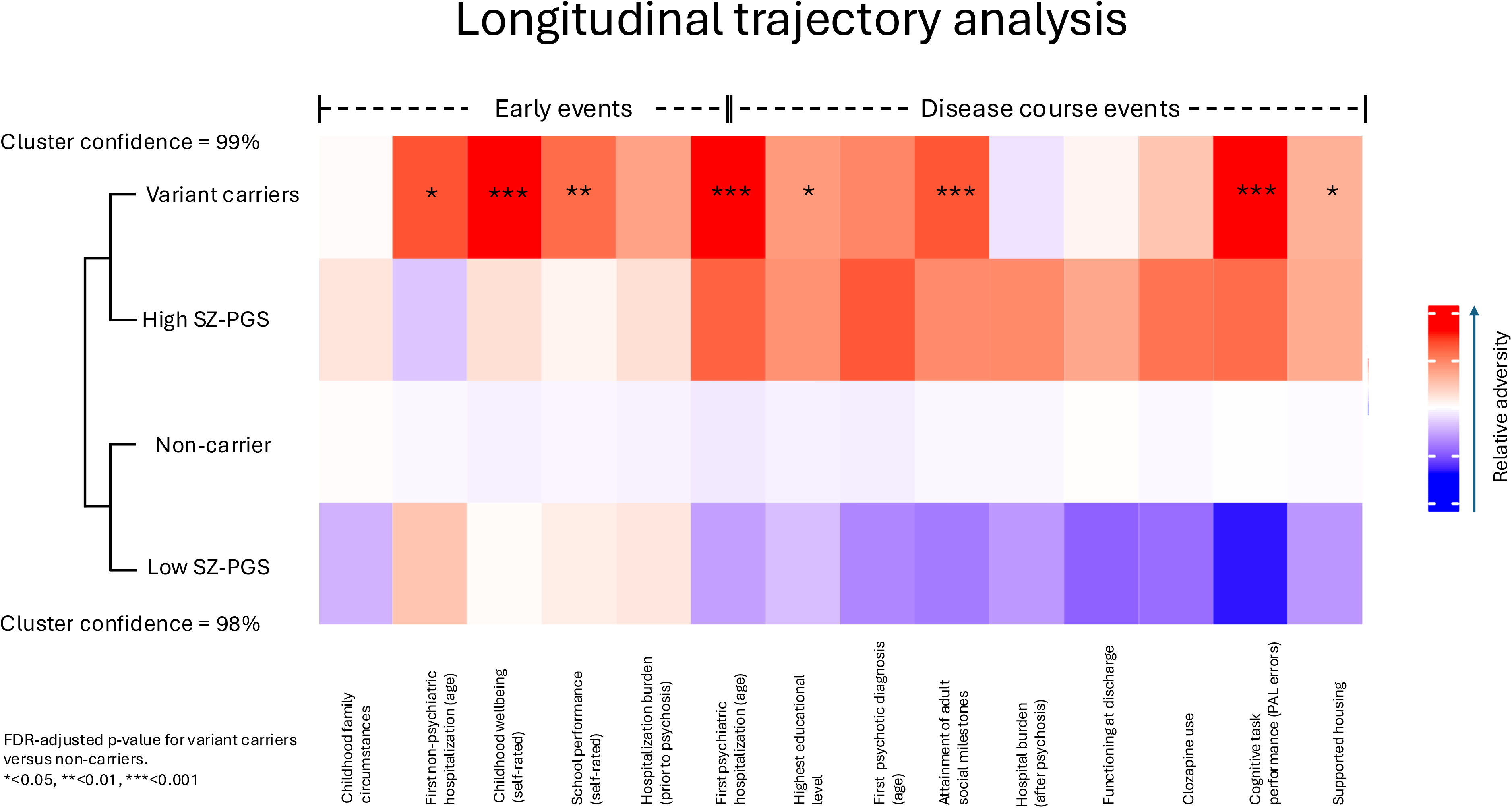
Longitudinal clinical and functional trajectories across genetic subgroups. Longitudinal clinical, functional, and neuropsychological measures across variant carriers, individuals with high or low SZ-PGS, and non-carriers. Measures span early-life adversity, hospital care, illness onset, adult functioning, and neuropsychological test performance. Warmer colors indicate relatively less favorable outcomes. The dendrogram shows the clustering of the genetic subgroups. Asterisks indicate FDR-adjusted significance between variant carriers and non-carriers (*p < 0.05, **p < 0.01, and ***p < 0.001).

#### Variant carriers had greater medical needs in childhood

The life-course analysis identified hospital admissions as one of the clearest early-life differences between variant carriers and non-carriers [**Fig. 3**], prompting us to examine both the timing and the diagnostic composition of these admissions in greater detail. First, we used time-to-event analyses to model age at first psychiatric and non-psychiatric hospital admission during childhood. We found no evidence that SZ-PGS was associated with childhood hospital admissions. Variant carriers, however, had earlier admissions for both primary psychiatric diagnoses (HR=2.30 [1.48-3.57], p=2.05e-04) and primary non-psychiatric diagnoses (HR=1.40 [1.10-1.79], p=6.88e-03) [**Fig. 4**]. The association with non-psychiatric hospitalization was strongest during the first five years of life (HR = 1.71 [1.26–2.33], p = 6.80e-04), while psychiatric hospitalizations were uncommon during this period and occurred more frequently during adolescence. Non-carriers with NDD variants showed a similar pattern, with earlier psychiatric and non-psychiatric admissions than other non-carriers (HR=2.11 [1.18-3.76], p=0.01; HR=1.87 [1.37-2.53], p=6.69e-05, respectively) [Fig. S5a-b]. In a sensitivity analysis we excluded all psychiatric hospitalizations due to ID/NDD diagnoses [Methods], but the association observed for variant carriers remained essentially unchanged (HR=2.28 [1.45-3.57], p=3.43e-04), whereas the association was no longer significant for non-carriers with NDD variants (HR=1.81 [0.96-3.40], p=0.066).

**Figure 4.**
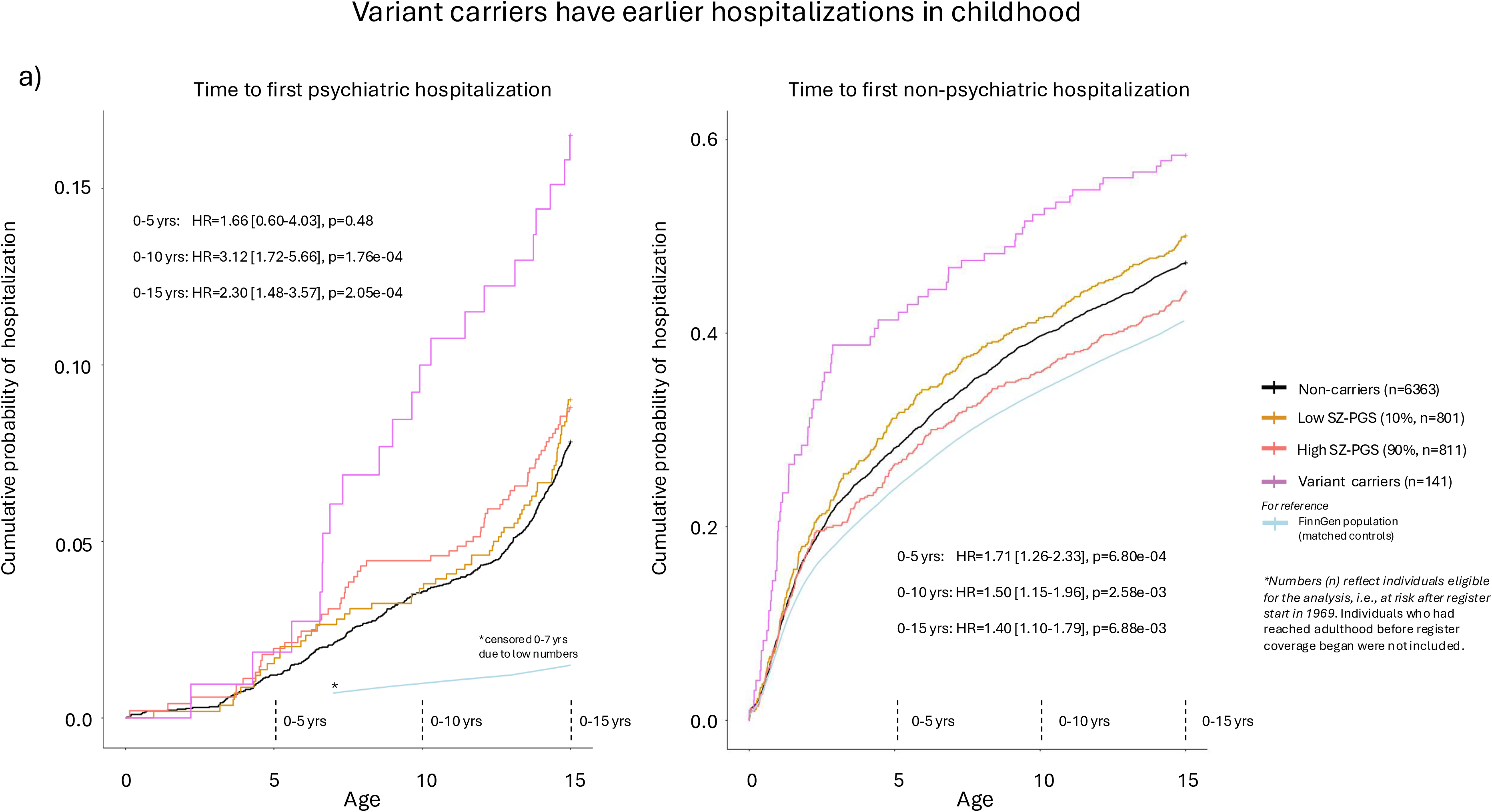
Earlier hospital admissions among variant carriers in childhood. Estimated cumulative probabilities of first psychiatric and non-psychiatric hospitalization before age 15 years among variant carriers, non-carriers, and non-carriers with high or low SZ-PGS. Analyses accounted for delayed entry resulting from hospital-register coverage beginning on January 1, 1969. Variant carriers had earlier hospital admissions for both psychiatric **(a)** and non-psychiatric diagnoses **(b)** compared with non-carriers. Hazard ratios are shown for the age intervals 0–5, 0–10, and 0–15 years. Matched FinnGen population controls are shown for reference.

Second, we characterized the diagnostic composition of these early-life hospitalizations. We focused the analysis on non-psychiatric admissions because they were substantially more common in childhood and covered a much wider range of diagnoses than psychiatric admissions. Specifically, we assessed the burden of the primary hospital diagnoses within each top-level ICD-10 category [Methods], focusing on the 10 most prevalent categories [Fig. S6a]. Using Monte Carlo resampling, we found that variant carriers had a higher-than-expected burden (FDR<0.05) of diagnoses categorized as ‘Symptoms, signs and abnormal clinical and laboratory findings, not elsewhere classified’ and ‘Certain infectious and parasitic diseases’ [Fig. S6b]. By contrast, non-carriers with NDD variants had a more clearly defined diagnostic pattern with a markedly higher burden of diagnoses categorized as ‘Diseases of the nervous system’. Comparisons between all subgroups can be viewed in Fig. S7a-g.

### Variant carriers showed poorer cognitive performance

The life-course analysis [**Fig. 3**] indicated that poorer cognitive performance was the most prominent difference between variant carriers and non-carriers in adulthood. We therefore asked whether this cognitive signal varied across specific tests and whether it was related to a diagnosis of SZ or co-occurring ID/NDD diagnoses. To address this, we examined cognitive performance data collected at study inclusion from the Paired Associates Learning (PAL) test of visual learning and memory and the Reaction Time (RTI) test, both included in the CANTAB suite and commonly used to measure neurocognitive function in SZ and related disorders (13) [Methods]. Overall, variant carriers had poorer visual memory and learning, reflected in more adjusted total errors on the PAL test, compared with non-carriers in SUPER-Finland (β = 0.41 [0.24-0.58], p=2.01e-06) [**Fig. 5a**]. Non-carriers with NDD variants also had poorer visual memory and learning, with an even larger point estimate for PAL errors (β = 0.60 [0.36-0.85], p=1.37e-06). We did not observe any difference in median RTI test performance between variant carriers and non-carriers (β= -0.01 [-0.19-0.16], p=0.88), whereas non-carriers with NDD variants had significantly slower reaction times (β = 0.37 [0.12-0.62], p=3.52e-03). Thus, the cognitive signal among variant carriers appeared to be most clearly reflected in visual memory and learning.

**Figure 5.**
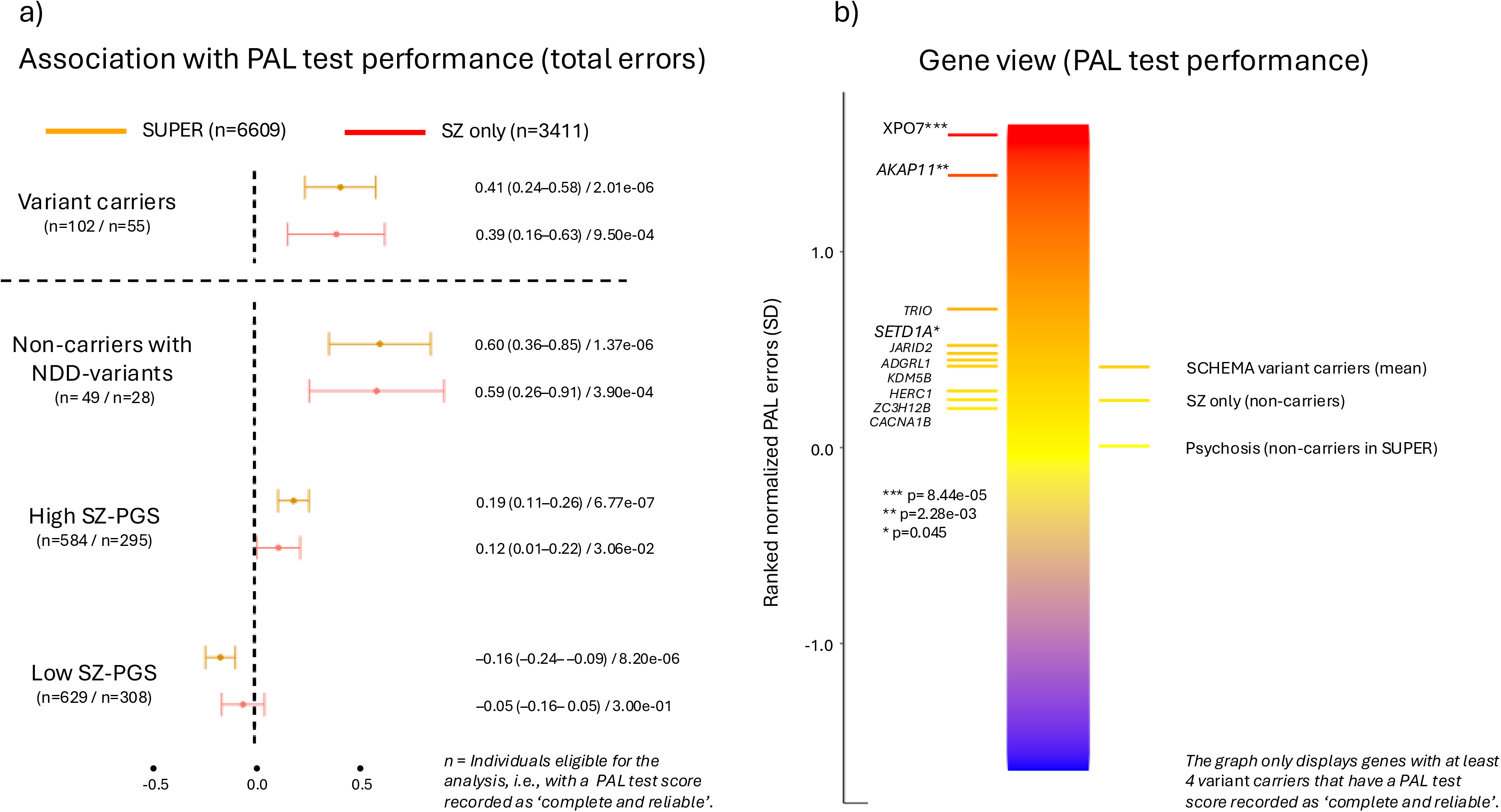
Variant carriers show poorer PAL test performance. **a)** Association between genetic subgroup status and PAL test performance in the full SUPER-Finland cohort and among individuals with a diagnosis of SZ. Points and horizontal lines indicate regression coefficients and 95% confidence intervals, respectively. **b)** Gene-level associations with PAL test performance among SZ-risk genes with at least four variant carriers whose PAL tests were rated as complete and reliable. Higher values indicate poorer performance. *XPO7* (p = 8.44e-05), *AKAP11* (p = 2.28e-03), and *SETD1A* (p = 0.045) showed individual associations with poorer PAL test performance.

To assess whether the poorer visual memory and learning of variant carriers reflected differences in diagnostic composition, we restricted the analysis to individuals with a diagnosis of SZ (n=5,479, of whom 3,411 had eligible PAL data, **Methods**). Within this subgroup, variant carriers continued to show poorer visual memory and learning than non-carriers (β = 0.39 [0.16-0.63], p=9.50e-04), whereas the association between SZ-PGS and PAL errors was attenuated [**Fig. 5a**]. This suggests that the SZ-PGS association observed in the full cohort partly reflected diagnostic composition. Among individuals with psychotic disorders other than SZ (n=4,342, of whom 3,198 had eligible PAL data), the association between variant carrier status and PAL errors also remained evident (β= 0.43 [0.19-0.67], p=4.63e-04).

Because poorer visual memory and learning could reflect co-occurring ID/NDD diagnoses, we examined register-recorded ID/NDD diagnoses [Methods]. These diagnoses were more common among variant carriers than among non-carriers (11.0% vs 5.6%; OR=1.99 [1.20-3.29], p = 7.8e-03), but not to the same extent as among non-carriers with NDD variants (19.5% vs 5.6%; OR=3.96 [2.29-6.83], p=8.03e-07).

However, after excluding all variant carriers with an ID/NDD diagnosis (n=18, 11.0%), the association between variant carrier status and PAL errors remained the same (β = 0.41 [0.24-0.59], p=3.74e-06). Furthermore, variant carriers whose affected SCHEMA-nominated gene had previously been associated with NDD (n=92, of whom 59 had eligible PAL data) had a slightly lower point estimate for PAL errors than variant carriers overall (β = 0.37 vs β = 0.41). Among variant carriers in the 11 SCHEMA-nominated genes previously reported to cause dominant NDD syndromes through haploinsufficiency (n=40, of whom 28 had eligible PAL data), a slightly higher point estimate for PAL errors was observed (β = 0.47), but this estimate was not detectably larger than that observed in other variant carriers (p = 0.67) [Fig. S8].

### Gene-level analyses of cognitive and diagnostic profiles

To examine gene-level associations with visual memory and learning, given the small number of variant carriers per gene, we focused our analysis on 10 SZ-risk genes. These were the genes with the largest numbers of variant carriers whose PAL tests had been rated by study personnel as “complete and reliable” (range = 4–12 variant carriers per gene) [Methods; **Fig. 5b**]. Among these genes, *XPO7* and *AKAP11* variant carriers showed the largest differences in PAL errors relative to non-carriers (*XPO7*: β = 1.71 [0.86-2.56], p=8.44e-05; *AKAP11*: β = 1.32 [0.47-2.17], p=2.28e-03). *SETD1A* variant carriers also showed a nominally significant association with poorer visual memory and learning compared to non-carriers (β= 0.65 [0.014-1.30], p=0.045). The gene-level PAL results were consistent in empirical resampling analyses (*XPO7*, empirical p < 1.0e-04; *AKAP11*, empirical p = 3.00e-03; *SETD1A*, empirical p = 0.046). Overall, 37.4% (n=61) of variant carriers did not have a PAL test rated as complete and reliable, which was slightly higher than among non-carriers (32.3%), but the difference was not significant (p=0.07).

#### Evidence of gene-specific phenotypic differences

To investigate whether psychiatric profiles differed across SZ-risk genes, we focused on the affective and non-affective disorder diagnostic dimensions, which are typically used to differentiate between diagnostic categories (14). BD and SZ broadly represent these two dimensions, and although they usually differ clinically, the disorders have overlapping genetic architectures, with both shared and disorder-specific risk loci (15-18). Because SZ (n=5,479) and BD (n=2,461) were the two most common psychotic diagnoses in the SUPER-Finland study (562 individuals had received both diagnoses), these were also the diagnostic dimensions for which we had the greatest power to detect differences. The SCHEMA analysis identified 7 SZ-associated genes that have also been associated with BD (*AKAP11*, *ATP9A*, *CUL1*, *HERC1*, *KDM5B, RB1CC1*, and *SP4*, n = 48 variant carriers) (10). We found that variant carriers in these 7 genes had a cumulative burden across all affective diagnoses [Methods] similar to that of non-carriers with a diagnosis of BD, and significantly higher than that of non-carriers with a SZ diagnosis [**Fig. 6a**]. At the same time, these variant carriers had a substantially higher burden of non-affective psychotic diagnoses than non-carriers with a diagnosis of BD (significance assessed by empirical resampling across 10,000 iterations) [**Fig. 6b**].

**Figure 6.**
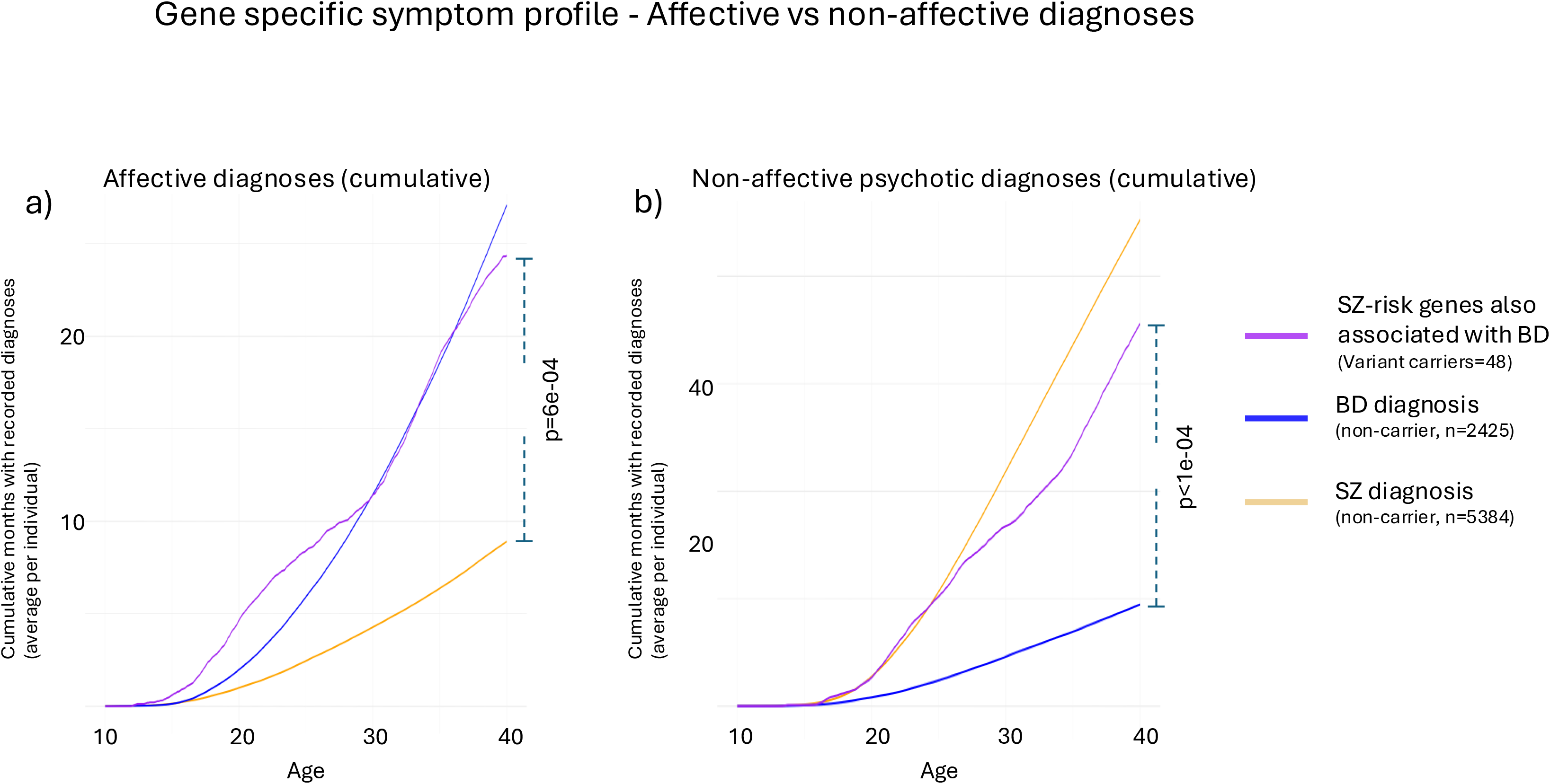
Gene-specific affective and non-affective symptom profiles. Cumulative registry-recorded affective diagnoses and non-affective psychotic diagnoses among variant carriers in the seven SZ-risk genes also associated with BD (n = 48), compared with non-carriers with a diagnosis of BD or SZ. **Panel a** shows the average cumulative affective diagnostic burden, and **panel b** shows the average cumulative non-affective psychotic diagnostic burden. Curves show the average cumulative number of recorded diagnoses per individual across age. P-values were estimated using resampling inference.

## Discussion

Here we studied the disease trajectories of individuals with psychotic disorders who carried damaging variants in SCHEMA-nominated SZ-risk genes associated with a many-fold increase in risk for SZ. Schizophrenia (SZ) and related psychotic disorders have overlapping, heterogeneous symptoms, with disease courses that can vary even between individuals with the same diagnosis (8, 19, 20). We combined the strength of broad recruitment across psychotic disorders in the SUPER-Finland study with decades of longitudinal medical register data and information from neuropsychological tests, questionnaires, and participant interviews (8, 11). Our findings provide new insights into how damaging variants in SZ-risk genes influence the course of psychotic disorders, highlighting associations with greater childhood adversity, earlier hospitalizations, and poorer cognitive performance. High-impact genetic markers for psychosis risk have remained scarce in psychiatric research. These high-risk variants provide new opportunities for studying the biological mechanisms underlying psychotic disorders because they directly implicate specific genes and often loss of function as the likely mode of effect. Understanding the phenotypic impact of these variants, including early-life features that precede psychotic illness onset, is important for interpreting their biological consequences and may ultimately improve the identification of individuals at risk and inform patient management.

The liability-threshold model proposes that a condition develops when the combined effects of an individual’s risk factors exceed a certain threshold (21, 22). In line with this model, variant carriers with a diagnosis of SZ had lower SZ-PGS than non-carriers with the same diagnosis, but higher SZ-PGS than population controls. This suggests that variant carriers may require a smaller polygenic contribution for psychotic illness to develop, while polygenic background still contributes to disease liability. This pattern was not observed among non-carriers with NDD variants, and the reduction in SZ-PGS was larger among variant carriers whose affected genes had the highest SCHEMA-reported SZ effect sizes, supporting a direct contribution of these damaging variants to disease liability. These findings extend previous reports on high-impact CNVs in SZ (23), which suggest that the polygenic background modulates penetrance, and are consistent with findings in autism spectrum disorders (24) and intellectual disability (12).

### Schizophrenia as a neurodevelopmental disorder

The widely accepted neurodevelopmental model of SZ posits that neurodevelopmental processes contributing to the disorder begin long before psychotic illness onset (25-27), consistent with evidence of premorbid deficits (28). The damaging variants examined here provide a genetic entry point for interrogating this model. Consistent with this model, differences between variant carriers and non-carriers were most apparent during early life, before the first psychotic disorder diagnosis. Variant carriers reported poorer childhood wellbeing and school performance and had earlier hospital admissions for both psychiatric and non-psychiatric causes. Support for gene–environment effects has been provided by previously reported associations between childhood trauma and genetic liability for SZ in population cohorts (29). However, in our within-case study, we did not detect associations between SZ-PGS and our childhood measures, and the group with the highest SZ-PGS primarily showed less favorable disease trajectories after illness onset. Our findings therefore add a rare-variant perspective in support of the neurodevelopmental model, linking damaging variants in SCHEMA-nominated SZ-risk genes to phenotypic differences detectable before the first recorded psychotic diagnosis.

During adulthood, variant carriers showed fewer differences from non-carriers, except for poorer visual memory and learning, as measured by the PAL test, which assesses cognitive functions relevant to psychotic disorders (13, 30-32). We observed that the association with PAL errors was seen across variant carriers, and not concentrated to variant carriers whose affected SCHEMA-nominated genes had previously been associated with NDD syndromes. Non-carriers with NDD variants also showed poorer visual memory and learning and earlier childhood hospitalizations. However, the diagnostic composition of their childhood hospitalizations differed from that of SCHEMA variant carriers. They also showed slower reaction times on the RTI test, whereas no such association was detected among SCHEMA variant carriers, suggesting a potentially different cognitive profile. Non-carriers with NDD variants and a diagnosis of SZ also had a SZ-PGS comparable to that of other non-carriers with SZ, implying that disruptive variants in haploinsufficient NDD genes not implicated by SCHEMA require a greater background polygenic contribution for psychosis to develop than damaging variants in SCHEMA-nominated genes. Similarly, among NFID participants ascertained for intellectual disability, only those with a psychotic diagnosis had a SZ-PGS similar to that of non-carriers in SUPER-Finland. Together, these findings suggest that, although the genetic architectures of psychosis and NDD syndromes overlap, similar neurodevelopmental phenotypes can be associated with different levels of genetic liability to psychosis.

### Variant effects can be observed across psychotic diagnoses

Predicting an individual’s disease course remains challenging because clinical features overlap across psychotic disorders, while simultaneously varying among individuals with the same diagnosis (8, 19). However, genetic findings could potentially provide relevant information beyond these diagnostic categories. The broad recruitment in the SUPER-Finland study [Methods] allowed us to interrogate the effects of the damaging variants across the psychotic spectrum. Our findings suggest that the phenotypic effects of the damaging SZ-associated variants were not confined to individuals with a diagnosis of SZ but extended across psychotic disorders. The association between variant carrier status and poorer visual memory and learning remained evident both among participants with SZ and among those with other psychotic disorders. The similar distribution of major psychotic disorder diagnoses among variant carriers and non-carriers also suggests that differences in diagnostic composition is unlikely to have been a major source of bias in our within-case comparisons. This is also important for the clinical interpretation of these variants, because it suggests that their penetrance for broader psychosis-spectrum and cognitive phenotypes may be greater than is captured by estimates based on SZ diagnosis alone. In support of this interpretation, a recent study in the UK Biobank also showed that rare damaging variants in SZ-risk genes identified by the 2022 SCHEMA analysis (5) were associated with poorer cognition among individuals from the general population without a psychotic diagnosis (33).

The results for the variant carriers contrasted with those observed among non-carriers at the tails of the SZ-PGS distribution. Non-carriers in the highest SZ-PGS decile had more PAL errors in SUPER-Finland, but the association was attenuated and only nominally significant when the analysis was restricted to individuals with a diagnosis of SZ. Individuals with a diagnosis of SZ performed markedly worse on the PAL test than those without a diagnosis of SZ, suggesting that the association between SZ-PGS and PAL performance may partly reflect the association between SZ-PGS and SZ diagnosis. In line with our results, a previous study of 3,034 individuals with a diagnosis of SZ found no significant association between SZ-PGS and estimated general intelligence (34). The prevalence of SZ differed markedly between the high and low SZ-PGS groups, but neither group showed corresponding differences in childhood outcomes. This suggests that SZ-PGS may primarily capture liability to SZ rather than early neurodevelopmental differences across the psychosis spectrum.

### Variant carriers showed evidence of gene-specific phenotypic profiles

Because carrier numbers were small, we treated variant carriers as a single group in many analyses. Nevertheless, the gene-level results suggest that the phenotypic effects of damaging SZ-associated variants may differ according to the affected gene. Although SCHEMA-reported gene-level effect sizes were associated with SZ-PGS [Fig. S2], phenotypic associations varied across genes and outcomes. In gene-level analyses, three genes stood out: *AKAP11*, *XPO7,* and *SETD1A*, for which gene-specific signals were detectable despite small carrier numbers. Interestingly, *XPO7* and *AKAP11* variant carriers had the largest point estimates for PAL errors, although neither gene is yet established as a cause of a monogenic NDD syndrome. Carriers of variants in *SETD1A*, a SCHEMA-nominated gene previously known to associate with NDD (35), also showed poorer visual memory and learning compared with non-carriers, but did not show detectably poorer performance than variant carriers overall. Variant carriers in these 3 genes had some of the lowest mean SZ-PGS values, and both *AKAP11* and *SETD1A* variant carriers had an average SZ-PGS close to that of population controls. Our analysis also revealed that variant carriers in SZ-risk genes that have independently been associated with BD (7 genes, 48 variant carriers) had a high cumulative burden of affective diagnoses, while simultaneously having a high burden of non-affective psychotic diagnoses. The results are consistent with previous evidence for shared common-and rare-variant liability between SZ and BD (15, 17) and illustrate how this shared liability can manifest in overlapping diagnostic histories. The results also highlight the limitations of a strict divide between affective and non-affective diagnostic dimensions.

### Study limitations

We recognize several limitations. First, the small number of variant carriers limited the resolution of many analyses, particularly those examining gene-specific and variant-specific effects. The variant carriers were therefore treated as a group in many analyses, despite the expected–and here observed– phenotypic heterogeneity across affected genes. Second, neuropsychological test performance was only assessed cross-sectionally at study inclusion, and interview, questionnaire, and register-based measures had to serve as proxies for cognitive function and wellbeing during childhood and early stages of illness. Furthermore, only PAL tests rated by study personnel as complete and reliable were included in the analysis [Methods]. We therefore likely did not capture the full range of possible test outcomes, potentially biasing the cognitive analyses toward higher-functioning participants and underestimating the magnitude of the cognitive differences. Finally, although Finnish healthcare registers are of high quality (36), register-based diagnoses are subject to variable classification due to local diagnostic traditions and changes in diagnostic practice over time (37). However, the study also has important strengths, particularly the recruitment of participants across psychotic disorders rather than exclusively among individuals with a diagnosis of SZ. This broad ascertainment enabled comparisons across diagnostic groups and the assessment of phenotypic features that would not have been observable in a cohort restricted to SZ.

## Conclusion

The disease trajectories of carriers of ultra-rare damaging variants in SZ-risk genes differed from those of non-carriers most prominently early in life, before the first recorded psychotic diagnosis. Variant carriers reported greater childhood adversity and had earlier hospital admissions but also had poorer neuropsychological test performance in adulthood. These differences were detectable across psychotic disorders and suggest that the phenotypic consequences of the SZ-associated variants extend beyond the categorical SZ diagnosis used to identify these risk genes. Variant carriers had lower SZ-PGS than non-carriers in SUPER-Finland, including among individuals with a diagnosis of SZ, supporting a liability-threshold model in which rare large-effect variants can contribute to crossing the threshold for psychosis at lower levels of common-variant liability. Exploratory gene-level analyses also suggested gene-specific variation in phenotypic profiles and polygenic liability to SZ. Together, these findings are consistent with the neurodevelopmental model of SZ and related psychotic disorders and highlight the potential value of integrating rare variant information into future clinical risk stratification models.

## Methods

### The SUPER-Finland study

The SUPER-Finland study is part of the Stanley Global Neuropsychiatric Genetics Initiative and in total includes 10,474 adults with a history of at least one episode of psychotic illness (ICD-10: F20–F29, F31, F32.3, F33.3). Participants were recruited nationwide in Finland between 2016 and 2018, typically through their treating services (inpatient, outpatient, or primary health care). At study inclusion, trained clinical staff conducted interviews and administered questionnaires covering health-related factors, current and past wellbeing, sociodemographic characteristics, and early-life information, including school performance. Qualified personnel also supervised the cognitive testing using the Cambridge Neuropsychological Test Automated Battery (CANTAB). The study was approved by the Ethics Committee of the Hospital District of Helsinki and Uusimaa (202/13/03/00/15), and all participants provided written informed consent. Further details are provided by *Lähteenvuo et al* (11) and at https://thl.fi/en/research-and-development/thl-biobank/for-researchers/sample-collections/super-study.

### Finnish healthcare registers

Information on specific diagnoses (ICD codes) and their dates of recording was retrieved from the nationwide Finnish National Care Register for Health Care and the Register of Primary Health Care Visits (https://thl.fi/en/statistics-and-data/data-and-services/register-descriptions). The clinical registry data include information from 1) the Hospital Discharge Registry from 1969 onward; 2) specialized outpatient clinics from 1998 onward and 3) primary healthcare units from 2011 onward [Supp. Appendix]. Because the Finnish healthcare registries have been shown to be very reliable (36), we only considered diagnoses recorded in the registers, and not self-reported diagnoses obtained at study inclusion. Because of the large age span in SUPER-Finland (median birth year = 1970, IQR = 1958–1982) we focused most longitudinal register-based analyses on the Hospital Discharge Register, for which coverage was more comparable across generations.

#### Main psychotic disorder diagnosis

For certain analyses, each participant was assigned a main lifetime psychotic diagnosis using the hierarchical diagnostic model included in the SUPER-Finland study protocol (7, 11). The model ranks the five major diagnostic categories as: 1) schizophrenia (SZ), n=5,479; 2) schizoaffective disorder (SAD), n=874 (2,124); 3) bipolar disorder (BD), n= 1,494 (2,461); 4) major depressive disorder with psychotic features (psychotic MDD), n = 507 (1,542); 5) Other unranked psychotic diagnosis (n=1,067). The numbers represent participants whose highest-ranked psychotic diagnosis was in that category, while the counts in parentheses represent participants who had received that diagnosis at any time. We did not consider self-reported diagnoses, and 400 participants had no psychotic diagnosis recorded in the available healthcare registers, meaning no main lifetime psychotic diagnosis was assigned. This diagnostic ranking, which is considered to reflect clinical severity, has also been shown to be valid in the SUPER-Finland study (8). For analyses encompassing the full SUPER-Finland cohort, the assigned main lifetime psychotic diagnosis was included as a covariate.

### Phenotypic and clinical endpoints

#### Longitudinal endpoints

Diagnostic endpoints were exclusively defined using healthcare register data. For each endpoint, age at first diagnosis was defined as the age at the first corresponding register diagnosis. Because a psychotic disorder was required for inclusion in the SUPER-Finland study, age at psychotic illness onset was broadly defined as age at the first registry-recorded diagnosis indicating a psychotic illness. Similarly, BD was defined to include all ICD codes for BD (ICD-10 equivalents: F30.2 and F31.X). Details on psychiatric hospitalization burden, a measure of the yearly need for psychiatric hospital care, have previously been published (8) and shown to be a valid measure of clinical severity.

For analyses of early-life hospital care, we considered age at the first hospital admission with either a psychiatric or non-psychiatric primary diagnosis. Age- and sex-matched population reference groups from the FinnGen study (38) were included to contextualize the hospitalization patterns observed in SUPER-Finland [Supp. Appendix, p. 4]. For non-psychiatric primary diagnoses, admission causes were grouped into broad ICD top-level diagnostic categories (e.g., “Infectious and parasitic diseases” included ICD-10 A- and B-codes). Diagnoses were harmonized at the broad diagnostic-category level across ICD-8, ICD-9, and ICD-10, and the analysis focused on the 10 most common non-psychiatric diagnostic categories up to 15 years of age. For each diagnostic category, cumulative hospitalization burden was calculated by summing age-specific mean inpatient days, using the number of participants contributing to each one-year age interval as the denominator. Variant carriers and non-carriers with NDD variants were compared with non-carriers using empirical Monte Carlo resampling. Full diagnostic definitions and category mappings are provided in the Supplementary Appendix [pp. 15].

Register-recorded intellectual disability and/or neurodevelopmental disorder (ID/NDD) diagnoses were defined using ICD codes corresponding to intellectual disability and autism spectrum disorders. Because register-based identification of ID/NDD is sensitive to the breadth of the diagnostic definition, we used a narrow definition based on diagnoses that could be interpreted consistently across ICD versions. This endpoint was used primarily in sensitivity analyses excluding participants with register-recorded ID/NDD diagnoses. Full endpoint definitions, including the ICD codes used, are provided in the Supplementary Appendix [p. 16].

#### Functioning level at discharge

Psychiatric wards in Finland have rated functioning level at discharge using the Global Assessment Scale (GAS) (39) since 1992. Each participant’s mean GAS score across all hospitalizations was then used as one measure of disease severity (8,634 individuals had at least one discharge with a recorded GAS score).

#### Cross-sectional endpoints collected at study inclusion

Self-reported endpoints and cognitive test results were obtained from interviews, questionnaires, and assessments conducted at study inclusion. For the life-course overview shown in **Fig. 3**, these measures were considered together with the register-based endpoints described above. Interview and questionnaire items were grouped according to the life period and broad construct they were intended to assess. Childhood domains captured family circumstances, self-reported childhood wellbeing, and self-reported school performance, whereas adult domains captured the attainment of adult social milestones (e.g., employment, marriage, and parenthood) and current everyday functioning. Conceptually related items were combined into composite variables to reduce redundancy and to represent broader aspects of functioning that might not be adequately captured by individual items. Composite scores were calculated by summing their component items. The purpose of this overview was to assess whether phenotypic differences between the genetic subgroups were concentrated within particular periods of life, rather than to identify which individual interview or questionnaire item most strongly distinguished the groups. Full item wording, coding, and composite-variable definitions are provided in the Supplementary Appendix (pp. 9–11).

The highest level of education (low, intermediate, high) was defined according to the Finnish Health 2000 general population survey (40) and was recorded during the interview or collected from the Finnish School Registry. At inclusion, participants completed selected components of the Cambridge Neuropsychological Test Automated Battery (CANTAB). Cognitive performance was primarily indexed by adjusted total errors on the Paired Associates Learning (PAL) test, which measures visual memory and new learning. Only PAL tests rated by study personnel as “complete and reliable” were included (n = 6,609). Because PAL error scores showed a strongly bimodal distribution, we applied a rank-based inverse normal transformation before analysis. Reaction Time (RTI) performance, measured as the median reaction time, was analyzed as a secondary cognitive endpoint, with higher values reflecting slower reaction time (n = 7,495 participants completed the test).

#### Affective and non-affective diagnostic burden

We used both inpatient hospital and outpatient register records to capture diagnostic profiles rather than only hospital-care severity. Records were classified into two diagnostic dimensions: affective diagnoses, comprising all mood (affective) disorders (ICD-10 equivalent: F30-F39) and schizoaffective disorder; and non-affective psychotic diagnoses, comprising schizophrenia-spectrum and other non-affective psychotic disorders but excluding schizoaffective disorder. Because some episodes spanned multiple months, records were expanded into calendar-month blocks. Cumulative burden within each dimension was quantified from the number of calendar months containing at least one corresponding diagnostic record. Exact ICD-code definitions and details of the month-blocking procedure are provided in the Supplementary Appendix (p. 17).

### Genetic data

#### Sequencing and genotyping data

From the 10,474 participants, sequencing data was available for 10,096 participants (WES, n = 9,212; WGS, n = 884). All study participants were also genotyped using the Illumina Global Screening Array. Samples with poor genotype quality, ancestral outliers, and samples that mismatched with the recorded sex were excluded. Imputation was performed using a Finnish-specific reference panel, SISu version 3, and only reliably imputed variants (INFO score >0.8) were included. After genomic data quality control, 9,821 participants were included in the analyses.

#### Construction of SZ-PGS and comparative groups

The SZ-PGS was constructed using MegaPRS (41), which has been shown to perform well for psychiatric traits compared with other advanced PGS methods (42), using summary statistics from the largest available GWAS of SZ (Psychiatric Genomics Consortium, n cases = 74,776) (3). For comparisons based on the tails of the distribution, non-carriers in the lowest and highest SZ-PGS deciles were defined using the 10th and 90th percentile cutoffs, respectively. SZ-PGS was standardized as a Z-score using the mean and standard deviation among the 7,463 population reference participants from the Health 2000 study (40, 43). To provide a comparison with individuals ascertained for intellectual disability, we additionally examined 1,139 participants with available SZ-PGS from the Northern Finland Intellectual Disability (NFID) study, including individuals with intellectual disability of unknown etiology (44).

#### Identification of damaging variants in SCHEMA-nominated genes and definition of genetic subgroups

The sequencing data were processed at the Broad Institute of Harvard and MIT using a best-practice workflow and annotated with the variant effect predictor (VEP) (45). For heterozygous single nucleotide variants (SNVs) and small indels, an allelic balance between 20% and 80% was required. Predicted loss-of-function variants comprised stop-gained, frameshift, and canonical splice-site variants classified as high confidence by LOFTEE (46). After quality control, we screened participants for coding high-impact variants in SCHEMA-nominated SZ-risk genes. The high-impact definition was centered on high-confidence predicted loss-of-function variants but also included a restricted set of missense variants predicted to be highly deleterious by two complementary methods: AlphaMissense pathogenicity score ≥0.98 and MPC score ≥3. These stringent missense criteria were used to identify missense variants predicted to have substantial functional consequences while avoiding broader inclusion of missense variation.

Individuals with deleterious variants in non-SCHEMA genes indicating dominant NDD syndromes (group: non-carriers with NDD variants) were identified using a strict gene set constructed through a combination of ClinGen’s curated haploinsufficiency scores (47), gnomAD constraint scores, pathogenic variants reported to ClinVar (48) and *de novo* variant enrichment in developmental disorders (49). Variant details are found in Table S5, and all subgroup definitions can be found in Supp. Appendix (pp. 5–7).

### Statistical inference

#### Regression models

Cox regression and linear/logistic regression were used as appropriate for the outcome. The regression models included sex, year of birth, genotyping batch, and the first 10 principal components as covariates. Analyses were additionally stratified by diagnosis of SZ when relevant and sufficiently powered; otherwise, models were adjusted for the main lifetime psychotic diagnosis.

Transformations and analysis-specific covariates were used where appropriate. For analyses of childhood hospitalizations Cox regression models with delayed entry were used. Participants entered follow-up at birth or at the start of Hospital Discharge Register coverage, whichever occurred later, and contributed risk time until their first relevant hospitalization or censoring. The proportional-hazards assumption was evaluated using scaled Schoenfeld residuals; because the association with variant carrier status varied over childhood, cumulative analyses were performed with follow-up ending at ages 5, 10, and 15 years. Analyses included participants with available data for the relevant variables; no phenotype data was imputed.

#### Resampling-based statistical inference

For analyses in which the distribution of the test statistic was not suitable for regression-based inference, we used empirical Monte Carlo resampling to estimate statistical significance. Because the pool of eligible non-carriers was more than 50-fold larger than the variant carrier group, we considered this approach appropriate. Within each Monte Carlo iteration, comparison groups were sampled without replacement from the eligible non-carriers. To compare effect sizes between groups, bootstrap confidence intervals were calculated by resampling with replacement. Unless otherwise specified, Monte Carlo and bootstrap analyses were based on 10,000 iterations. Where applicable, p-values were adjusted for multiple testing using the Benjamini–Hochberg procedure, and results were considered statistically significant at FDR <0.05. For gene-level analyses with small carrier numbers, we also calculated empirical p-values by repeatedly sampling non-carrier groups and refitting the same covariate-adjusted regression model. All statistical analyses and figures were generated using R version 4.4.0.

## Data Availability

The individual-level data underlying this study cannot be shared or transferred outside the secure data environment of the University of Helsinki because they contain sensitive personal information. Aggregated data can be made available upon reasonable request to the authors.

## Additional Information

### Author contributions

A.K., O.P., M.J.D., B.M.N., and A.P. conceptualized the study and contributed to the analytic strategy. A.K. performed the statistical analyses and drafted the manuscript. J.Se., L.U., C.L., and E.H. contributed to data generation. J.Se., E.J.G., C.L., E.V., J.Su., M.L., T.P., O.K., A.T-H., J.L., E.I., T.Kie., K.L. and T.Kii. contributed to the interpretation of the data and analytical findings. All authors contributed to the interpretation of the findings and revision of the manuscript and approved its final version.

## Acknowledgements

**SUPER-Finland:** We want to thank the SUPER-Finland study participants. In addition, we want to thank the study nurses Panu Aunola, Marihelena Finne, Aino-Maija Haapasalo, Susanna Hotakainen, Ansa Järvinen, Veera Kemppainen, Kimmo Kontiainen, Maarit Kostiander, Eveliina Lehtinen, Maarit Lehtinen, Pia Maijala, Ulla Miettinen, Arja Minkkinen, Juha Mäkelä, Lea Nevalainen, Reijo Nevalainen, Paula Nurmi, Tero Rajatie, Outi Timonen, Kati Tuohimaa, Ulla Tyyni, Carita Vaittinen and Pia Virtanen, who worked in the field during the collection phase and the THL data collection and sample processing team whose work allowed for almost real-time surveillance of the collected material. Especially, we would like to thank Hannu Turunen for data management and Auli Toivola and Noora Ristiluoma for research coordination. We want to thank the outpatient policlinics and other healthcare units for their cooperation during the recruitment. We acknowledge the Genomics Platform of the Broad Institute of MIT and Harvard for genotyping the SUPER-Finland study samples.

**FinnGen:** We want to acknowledge the participants and investigators of the FinnGen study. The FinnGen project is funded by two grants from Business Finland (HUS 4685/31/2016 and UH 4386/31/2016) and the following industry partners: AbbVie Inc., Alnylam Pharmaceuticals, Inc., AstraZeneca UK Ltd, Bayer AG, Biogen MA Inc., Boehringer Ingelheim International GmbH, Bristol Myers Squibb Inc. (and Celgene Corporation C Celgene International II Sàrl), Genentech Inc., GlaxoSmithKline Intellectual Property Development Ltd., JohnsonCJohnson Innovative Medicine Inc., Maze Therapeutics Inc., Merck Sharp C Dohme LCC, Novartis AG, Pfizer Inc. and Sanofi US Services Inc.

## References

1. Kahn RS, Sommer IE, Murray RM, Meyer-Lindenberg A, Weinberger DR, Cannon TD, et al. Schizophrenia. Nature Reviews Disease Primers. 2015;1(1):15067.

2. Hilker R, Helenius D, Fagerlund B, Skytthe A, Christensen K, Werge TM, et al. Heritability of Schizophrenia and Schizophrenia Spectrum Based on the Nationwide Danish Twin Register. Biol Psychiatry. 2018;83(6):492–8.

3. Trubetskoy V, Pardiñas AF, Qi T, Panagiotaropoulou G, Awasthi S, Bigdeli TB, et al. Mapping genomic loci implicates genes and synaptic biology in schizophrenia. Nature. 2022;604(7906):502–8.

4. Marshall CR, Howrigan DP, Merico D, Thiruvahindrapuram B, Wu W, Greer DS, et al. Contribution of copy number variants to schizophrenia from a genome-wide study of 41,321 subjects. Nat Genet. 2017;49(1):27–35.

5. Singh T, Poterba T, Curtis D, Akil H, Al Eissa M, Barchas JD, et al. Rare coding variants in ten genes confer substantial risk for schizophrenia. Nature. 2022;604(7906):509–16.

6. Genovese G, Fromer M, Stahl EA, Ruderfer DM, Chambert K, Landén M, et al. Increased burden of ultra-rare protein-altering variants among 4,877 individuals with schizophrenia. Nat Neurosci. 2016;19(11):1433–41.

7. Ahti J, Kieseppä T, Suvisaari J, Suokas K, Holm M, Wegelius A, et al. Differences in psychosocial functioning between psychotic disorders in the Finnish SUPER study. Schizophrenia Research. 2022;244:10–7.

8. Kämpe A, Suvisaari J, Lähteenvuo M, Singh T, Ahola-Olli A, Urpa L, et al. Genetic contribution to disease-course severity and progression in the SUPER-Finland study, a cohort of 10,403 individuals with psychotic disorders. Mol Psychiatry. 2024.

9. Giangrande EJ, Kämpe A, Suvisaari J, Lähteenvuo M, Vartiainen E, Salo K, et al. Polygenic Scores for Schizophrenia and Educational Attainment Predict Global Functioning Across Psychiatric Hospitalization Among People with Schizophrenia. medRxiv. 2025:2025.05.20.25328039.

10. Sealock JM, Dowd C, Liao C, Ye R, Howrigan D, Satterstrom FK, et al. Analysis of Rare Coding Variation Identifies New Genetic Contributors to Schizophrenia. bioRxiv. 2026:2026.09.18.752670.

11. Lähteenvuo M, Ahola-Olli A, Suokas K, Holm M, Misiewicz Z, Jukuri T, et al. Cohort profile: SUPER-Finland – the Finnish study for hereditary mechanisms of psychotic disorders. BMJ Open. 2023;13(4):e070710.

12. Urpa L, Kurki MI, Rahikkala E, Hämäläinen E, Salomaa V, Suvisaari J, et al. Evidence for the additivity of rare and common variant burden throughout the spectrum of intellectual disability. Eur J Hum Genet. 2024;32(5):576–83.

13. Levaux M-N, Potvin S, Sepehry AA, Sablier J, Mendrek A, Stip E. Computerized assessment of cognition in schizophrenia: Promises and pitfalls of CANTAB. European Psychiatry. 2007;22(2):104–15.

14. Cerqueira RO, Ziebold C, Cavalcante D, Oliveira G, Vásquez J, Undurraga J, et al. Differences of affective and non-affective psychoses in early intervention services from Latin America. Journal of Affective Disorders. 2022;316:83–90.

15. Ruderfer DM, Ripke S, McQuillin A, Boocock J, Stahl EA, Pavlides JMW, et al. Genomic Dissection of Bipolar Disorder and Schizophrenia, Including 28 Subphenotypes. Cell. 2018;173(7):1705-15.e16.

16. Richards AL, Cardno A, Harold G, Craddock NJ, Di Florio A, Jones L, et al. Genetic Liabilities Differentiating Bipolar Disorder, Schizophrenia, and Major Depressive Disorder, and Phenotypic Heterogeneity in Bipolar Disorder. JAMA Psychiatry. 2022;79(10):1032–9.

17. Palmer DS, Howrigan DP, Chapman SB, Adolfsson R, Bass N, Blackwood D, et al. Exome sequencing in bipolar disorder identifies AKAP11 as a risk gene shared with schizophrenia. Nat Genet. 2022;54(5):541–7.

18. Thorgeirsson TE, Tragante V, Sveinbjornsson G, Jonsdottir GA, Walters GB, Ivarsdottir EV, et al. Rare loss-of-function variants in HECTD2 and AKAP11 confer risk of bipolar disorder. Nature Genetics. 2025;57(4):851–5.

19. Bourque V-R, Poulain C, Proulx C, Moreau CA, Joober R, Forgeot d’Arc B, et al. Genetic and phenotypic similarity across major psychiatric disorders: a systematic review and quantitative assessment. Translational Psychiatry. 2024;14(1):171.

20. Jääskeläinen E, Juola P, Hirvonen N, McGrath JJ, Saha S, Isohanni M, et al. A systematic review and meta-analysis of recovery in schizophrenia. Schizophr Bull. 2013;39(6):1296–306.

21. Falconer DS. The inheritance of liability to diseases with variable age of onset, with particular reference to diabetes mellitus. Annals of Human Genetics. 1967;31(1):1–20.

22. Kämpe A, Blomqvist Picard O, Eisfeldt J, Lindstrand A. Moving beyond monogenic disorders in clinical healthcare. Nature Biotechnology. 2026;44(1):21–5.

23. Bergen SE, Ploner A, Howrigan D, O’Donovan MC, Smoller JW, Sullivan PF, et al. Joint Contributions of Rare Copy Number Variants and Common SNPs to Risk for Schizophrenia. Am J Psychiatry. 2019;176(1):29–35.

24. Antaki D, Guevara J, Maihofer AX, Klein M, Gujral M, Grove J, et al. A phenotypic spectrum of autism is attributable to the combined effects of rare variants, polygenic risk and sex. Nature Genetics. 2022;54(9):1284–92.

25. Nagendra A, Mesholam-Gately R, Shafrin J, Keshavan MS. Reframing schizophrenia as a neurodevelopmental syndrome: The scientific and social imperative. Schizophrenia Research. 2026;292:96–102.

26. Rapoport JL, Giedd JN, Gogtay N. Neurodevelopmental model of schizophrenia: update 2012. Mol Psychiatry. 2012;17(12):1228–38.

27. Chien Y-L, Lin H-Y, Tung Y-H, Hwang T-J, Chen C-L, Wu C-S, et al. Neurodevelopmental model of schizophrenia revisited: similarity in individual deviation and idiosyncrasy from the normative model of whole-brain white matter tracts and shared brain-cognition covariation with ADHD and ASD. Molecular Psychiatry. 2022;27(8):3262–71.

28. Mohn-Haugen CR, Mohn C, Larøi F, Teigset CM, Øie MG, Rund BR. A systematic review of premorbid cognitive functioning and its timing of onset in schizophrenia spectrum disorders. Schizophrenia Research: Cognition. 2022;28:100246.

29. Sallis HM, Croft J, Havdahl A, Jones HJ, Dunn EC, Davey Smith G, et al. Genetic liability to schizophrenia is associated with exposure to traumatic events in childhood. Psychol Med. 2021;51(11):1814–21.

30. Junkkila J, Oja S, Laine M, Karrasch M. Applicability of the CANTAB-PAL computerized memory test in identifying amnestic mild cognitive impairment and Alzheimer’s disease. Dement Geriatr Cogn Disord. 2012;34(2):83–9.

31. Campos-Magdaleno M, Leiva D, Pereiro AX, Lojo-Seoane C, Mallo SC, Facal D, et al. Changes in visual memory in mild cognitive impairment: a longitudinal study with CANTAB. Psychol Med. 2021;51(14):2465–75.

32. Ashford MT, Aaronson A, Kwang W, Eichenbaum J, Gummadi S, Jin C, et al. Unsupervised Online Paired Associates Learning Task from the Cambridge Neuropsychological Test Automated Battery (CANTAB®) in the Brain Health Registry. The Journal of Prevention of Alzheimer’s Disease. 2024;11(2):514–24.

33. Fenner E, Holmans P, O’Donovan MC, Owen MJ, Walters JTR, Rees E. Analysis of rare coding variants in schizophrenia-associated genes and generalised cognition in the UK Biobank. Molecular Psychiatry. 2026.

34. Richards AL, Pardiñas AF, Frizzati A, Tansey KE, Lynham AJ, Holmans P, et al. The Relationship Between Polygenic Risk Scores and Cognition in Schizophrenia. Schizophr Bull. 2020;46(2):336–44.

35. Singh T, Kurki MI, Curtis D, Purcell SM, Crooks L, McRae J, et al. Rare loss-of-function variants in SETD1A are associated with schizophrenia and developmental disorders. Nat Neurosci. 2016;19(4):571–7.

36. Sund R. Quality of the Finnish Hospital Discharge Register: a systematic review. Scand J Public Health. 2012;40(6):505–15.

37. Nordgaard J, Jessen K, Sæbye D, Parnas J. Variability in clinical diagnoses during the ICD-8 and ICD-10 era. Soc Psychiatry Psychiatr Epidemiol. 2016;51(9):1293–9.

38. Kurki MI, Karjalainen J, Palta P, Sipilä TP, Kristiansson K, Donner KM, et al. FinnGen provides genetic insights from a well-phenotyped isolated population. Nature. 2023;613(7944):508–18.

39. Endicott J, Spitzer RL, Fleiss JL, Cohen J. The Global Assessment Scale: A Procedure for Measuring Overall Severity of Psychiatric Disturbance. Archives of General Psychiatry. 1976;33(6):766–71.

40. Aromaa AJ, Koskinen S, editors. HEALTH AND FUNCTIONAL CAPACITY IN FINLAND Baseline Results of the Health 2000 Health Examination Survey2004.

41. Zhang Q, Privé F, Vilhjálmsson B, Speed D. Improved genetic prediction of complex traits from individual-level data or summary statistics. Nature Communications. 2021;12(1):4192.

42. Ni G, Zeng J, Revez JA, Wang Y, Zheng Z, Ge T, et al. A Comparison of Ten Polygenic Score Methods for Psychiatric Disorders Applied Across Multiple Cohorts. Biol Psychiatry. 2021;90(9):611–20.

43. Health 2000. Data set description (THL): https://thl.fi/en/our-services/thl-biobank1/thl-biobank-datasets-and-samples.

44. Kurki MI, Saarentaus E, Pietiläinen O, Gormley P, Lal D, Kerminen S, et al. Contribution of rare and common variants to intellectual disability in a sub-isolate of Northern Finland. Nature Communications. 2019;10(1):410.

45. McLaren W, Gil L, Hunt SE, Riat HS, Ritchie GR, Thormann A, et al. The Ensembl Variant Effect Predictor. Genome Biol. 2016;17(1):122.

46. Karczewski KJ, Francioli LC, Tiao G, Cummings BB, Alföldi J, Wang Q, et al. The mutational constraint spectrum quantified from variation in 141,456 humans. Nature. 2020;581(7809):434–43.

47. Rehm HL, Berg JS, Brooks LD, Bustamante CD, Evans JP, Landrum MJ, et al. ClinGen — The Clinical Genome Resource. New England Journal of Medicine. 2015;372(23):2235–42.

48. Landrum MJ, Lee JM, Benson M, Brown GR, Chao C, Chitipiralla S, et al. ClinVar: improving access to variant interpretations and supporting evidence. Nucleic Acids Research. 2017;46(D1):D1062–D7.

49. Kaplanis J, Samocha KE, Wiel L, Zhang Z, Arvai KJ, Eberhardt RY, et al. Evidence for 28 genetic disorders discovered by combining healthcare and research data. Nature. 2020;586(7831):757–62.

