## Supplementary Appendix for "Ultra-rare coding variants in schizophrenia risk genes are associated with longitudinal phenotypic differences across psychotic disorders"

**INDEX**

### National healthcare registers in Finland

Finland has a long tradition of nationwide healthcare registers that can be linked at the individual level using personal identity codes. The Finnish Hospital Discharge Register has been available since 1969, providing five decades of follow-up. For the present study, diagnostic information was retrieved from the Finnish National Care Register for Health Care and the Register of Primary Health Care Visits. Together, these registers include inpatient hospital discharges since 1969, specialized outpatient visits since 1998, and primary healthcare visits since 2011.
The Finnish Hospital Discharge Register is one of the oldest nationwide individual-level hospital discharge registers in the world and has been extensively used in epidemiological and clinical research. It contains information on admission and discharge dates, hospital identifiers, and diagnoses recorded during hospital care. Diagnostic coding has followed the ICD system, with ICD-8 used during 1969–1986, ICD-9 during 1987–1995, and ICD-10 from 1996 onward. Because specialized outpatient care and primary healthcare data became available later, analyses intended to compare individuals across generations primarily relied on hospital discharge data.

The quality of the Finnish Hospital Discharge Register has been systematically examined by Sund et al (1), and the completeness and accuracy were judged to range from satisfactory to very good, and more than 95% of true hospital discharges could be identified in the register. These features make the Finnish registers particularly valuable for studying longitudinal disease trajectories.

**Register descriptions:** Nationwide Finnish National Care Register for Health Care and the Register of Primary Health Care Visits*:* https://thl.fi/en/statistics-and-data/data-and-services/register-descriptions

### The SUPER-Finland study

SUPER-Finland is a nationwide Finnish study within the Stanley Global Neuropsychiatric Genetics Initiative (https://www.broadinstitute.org/stanley-center-psychiatric-research/stanley-global). The cohort includes 10,474 consenting adults with a history of at least one psychotic episode. Recruitment was designed to capture the broad psychosis spectrum, including schizophrenia-spectrum psychotic disorders (ICD-10: F20, F22–F29), bipolar I disorder with psychosis (F30.2, F31.2, F31.5), and major depressive disorder with psychotic features (F32.3, F33.3). Participants were recruited from psychiatric inpatient and outpatient services, as well as from primary healthcare services, across all Finnish hospital districts. Recruitment was conducted by study psychiatrists and psychiatric nurses employed as part of the study staff, with the aim of obtaining a geographically broad and representative sample of individuals with psychotic disorders in Finland [Fig. SA1].

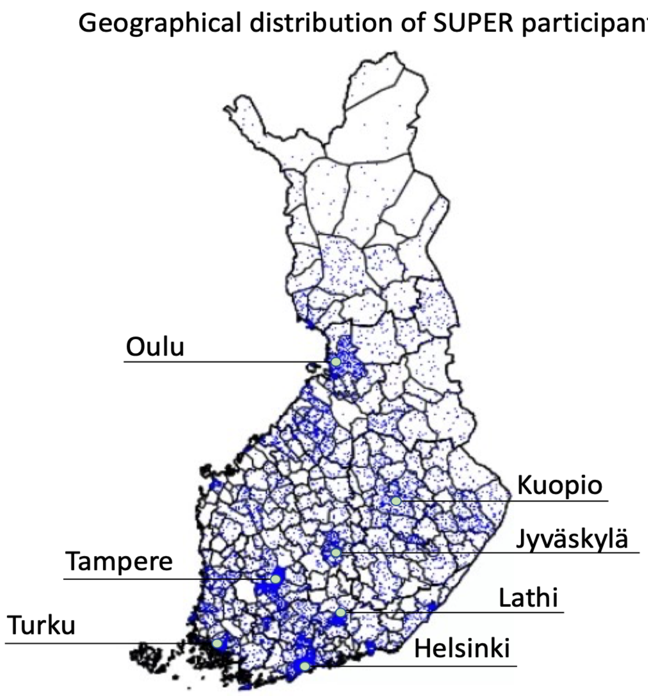

**Fig. SA1. Geographical distribution of SUPER-Finland participants.** Each blue dot represents one study participant. The seven largest urban areas in Finland are marked.
At study inclusion, participants underwent an in-person, protocol-guided interview conducted by study personnel. The assessment included interviews and questionnaires, as well as cognitive testing with selected tests from the Cambridge Neuropsychological Test Automated Battery (CANTAB).

Further information is available at the study website:
https://thl.fi/en/web/thl-biobank/for-researchers/sample-collections/super-study

### The FinnGen study

The FinnGen study includes 520,210 individuals from the Finnish population (data freeze 13; https://www.finngen.fi/en). FinnGen is a nationwide public–private partnership involving Finnish universities, university hospitals, national biobanks, and 13 international pharmaceutical companies, with linked nationwide healthcare register data for each participant (2). The overall study protocol was approved by the Coordinating Ethics Committee of the Hospital District of Helsinki and Uusimaa (HUS/990/2017).

Here, FinnGen was used only as a population reference for the analyses of time to first psychiatric and non-psychiatric hospitalization during childhood presented in **Fig. 4** of the main manuscript. The FinnGen reference groups were included to provide population context for the hospitalization patterns observed among variant carriers and the other genetic subgroups in SUPER-Finland.

**1. Time to first psychiatric hospitalization.** For the analysis in **Fig. 4a**, we used FinnGen to create an age- and sex-matched population reference group, excluding individuals with a psychotic disorder. This allowed us to estimate the need for psychiatric hospital care during childhood among individuals without a recorded psychotic disorder. The FinnGen reference group included 111,509 individuals with available register data.

Exclusion criteria: All individuals with a psychotic diagnosis. ICD codes captured by this regular expression:
^F2[0-9]|^F302|^F312|^F315|^F3[2-3]3|^295[0-9]|^296[1-4]E|^29[7-8][0-9][A-Z]|^29[7-9][0-9][0-9]

**2. Time to first non-psychiatric hospitalization.** For the analysis in **Fig. 4b,** we created an age- and sex-matched population reference group in FinnGen, excluding individuals who had been hospitalized with a psychiatric primary diagnosis or treated for a psychiatric diagnosis at a specialized psychiatric outpatient clinic. These criteria were used to estimate the need for non-psychiatric hospital care among individuals without recorded psychiatric hospitalizations or specialized outpatient psychiatric care. The FinnGen reference group included 77,193 individuals.

Exclusion criteria: Any hospital admission with a psychiatric primary diagnosis (ICD-10 F-category or equivalent) or treatment for a psychiatric diagnosis at a specialized psychiatric outpatient clinic.

**Genetic subgroups used to contextualize findings among variant carriers in SZ-risk genes.**

To understand how findings among variant carriers compared with those among other individuals with psychotic disorders in SUPER-Finland, we assigned participants to five genetic subgroups. In addition to variant carriers, these comprised a reference group of non-carriers, individuals in the highest (≥90th percentile) and lowest (≤10th percentile) SZ-PGS deciles, and individuals carrying disruptive variants in genes associated with dominant NDD syndromes (n = 87), here termed "non-carriers with NDD variants" [details below]. The NDD comparison group was included because several SCHEMA-nominated SZ-risk genes have also been implicated in NDDs and to assess whether the phenotypic patterns observed among variant carriers reflected broader consequences of disruptive variants associated with neurodevelopmental impairment. The five subgroup definitions are summarized in Table SA1.

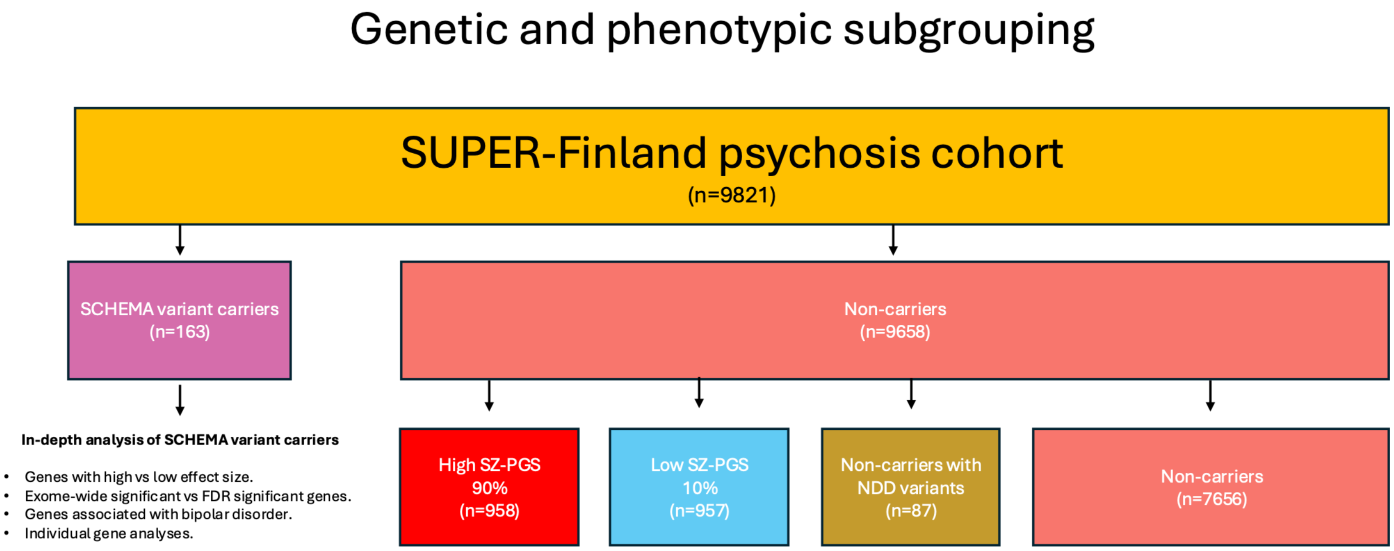

**Fig. SA2a.** Overview of the analyzed subgroups in SUPER-Finland. Flowchart showing how the genetic and phenotypic subgroups were defined. Depending on the analysis, comparisons were based on different levels of the hierarchy. Detailed subgroup definitions are provided below (p. 6).

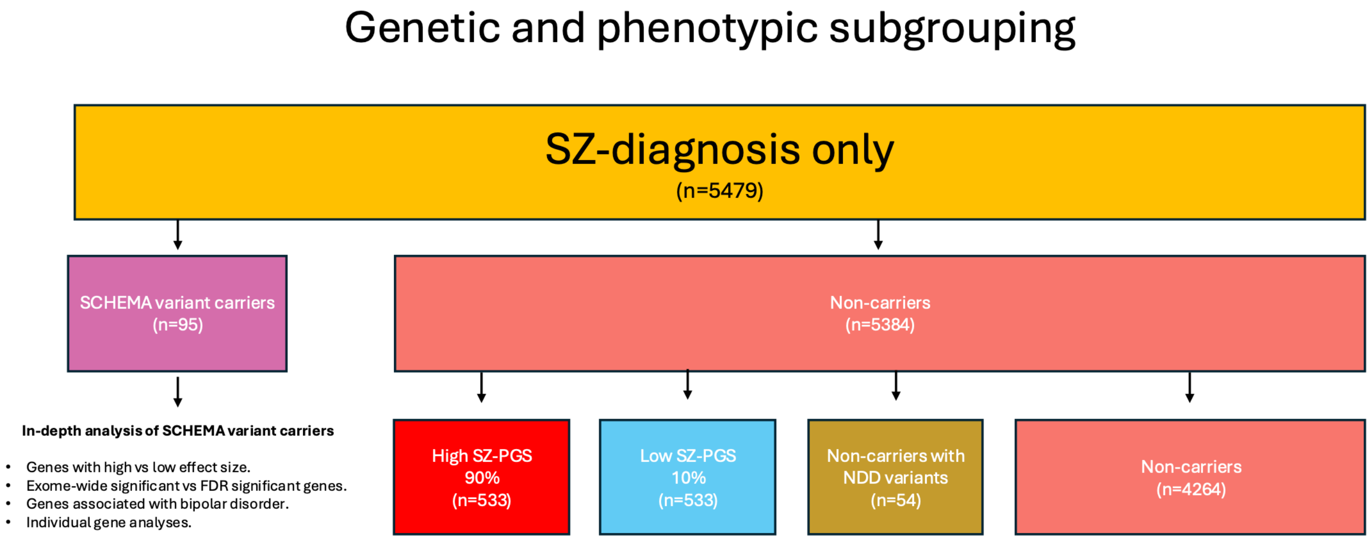

**Fig. SA2b.** Overview of the analyzed subgroups in SUPER-Finland: participants with SZ. Some analyses were restricted to SUPER-Finland participants with a diagnosis of SZ (n = 5,479). The high and low SZ-PGS groups were defined using SZ-PGS deciles calculated within this subset. Detailed definitions of all subgroups are provided below (p. 7).

**Table SA1. Detailed definitions of the genetic subgroups analyzed in SUPER-Finland.**

| | |
|---|---|
| 1 | Reference group (n = 7656): Non-carriers with an intermediate SZ-PGS (>10$^{th}$ to <90$^{th}$ percentile) |
| 2 | Variant carriers (n=163). Individuals harbouring damaging variants in SCHEMA-nominated SZ-risk genes. |
| 3 | Non-carriers with NDD variants (n=87): Individuals with a disruptive variant in a conservatively curated list of haploinsufficient genes known to cause dominant NDD syndromes. Variant details can be found in Table S5. |
| 4 | High SZ-PGS (n = 958): Non-carriers with a high SZ-PGS at or above the 90$^{th}$ percentile. |
| 5 | Low SZ-PGS (n = 957): Non-carriers with a low SZ-PGS at or below the 10$^{th}$ percentile. |

*Total n (SUPER-Finland) = 9821, not including ancestral outliers or samples that failed quality control.

#### Detailed subgroup definitions

**Variant carriers (n = 163).** This group comprised carriers of damaging coding variants in SZ-associated genes identified by the SCHEMA Consortium. The definition was centered on high-confidence predicted loss-of-function (pLoF) variants annotated by LOFTEE but also included a restricted set of likely deleterious missense variants that met both an MPC score ≥3 and an AlphaMissense pathogenicity score ≥0.98.

**SZ-PGS tail groups (high SZ-PGS, n = 958; low SZ-PGS, n = 957).** Summary statistics of the latest Psychiatric genomics consortium (PGC) schizophrenia GWAS (3) were used to calculate the SZ-PGS. Low and high SZ-PGS were defined as values within the lowest and highest deciles, respectively. For analyses restricted to participants with SZ, the decile cutoffs were recalculated within the SZ subset; otherwise they were calculated within the full SUPER-Finland cohort.

**Non-carriers with NDD variants (n=87):** To construct a comparison group with rare-variant liability for neurodevelopmental phenotypes, we identified individuals carrying variants associated with dominant NDD and/or intellectual disability (ID) syndromes. We restricted this group to variants expected to have high penetrance. We first identified 292 non-SCHEMA genes reported by Kaplanis et al. (4) to be significantly enriched for de novo variants in developmental disorders, based on more than 30,000 trios. This gene set was intersected with the Genomics England PanelApp curated ID gene panel (5), yielding 262 genes of interest.

Among these 262 genes, we first identified SUPER-Finland participants carrying disease-causing variants reported in ClinVar, the clinical variant database maintained by the National Center for Biotechnology Information (n = 44) (6). Only ClinVar variants classified as pathogenic or likely pathogenic, without conflicting interpretations, were included. To identify qualifying variants not reported in ClinVar, the 262 genes were further filtered using 1) gnomAD v4 gene-constraint scores (pLI >0.9) and 2) ClinGen's curated list of haploinsufficient genes (7). Carriers of loss-of-function variants in genes meeting these additional criteria were also included. LOFTEE (8) was used to filter out loss-of-function variants with flags indicating annotation uncertainties. We also excluded all variants with an allele count >10 in gnomAD v4. In total, 87 SUPER-Finland individuals were identified with disruptive variants in 54 different genes [Table S5], highly indicative of an NDD phenotype.

**Non-carriers (n=7656).** All study participants not included in any of the above subgroups were considered to belong to the group called 'non-carriers'. In analyses comparing the five mutually exclusive genetic subgroups, participants in the high and low SZ-PGS groups and participants with NDD variants were separated from the reference group, leaving 7,656 reference non-carriers with intermediate SZ-PGS. In analyses not specifically comparing these subgroups, participants in the SZ-PGS tails remained part of the broader non-carrier group, as specified for each analysis.

**Group ranking.** The five subgroups were mutually exclusive. Participants meeting the criteria for more than one subgroup were assigned to the highest-ranking applicable subgroup in the order shown in Table SA1.

### Life-course analyses

By combining the extensive interview and questionnaire data collected in SUPER-Finland with longitudinal Finnish healthcare register data, we were able to characterize participants' reported and register-recorded histories across different periods of life. The register data provided information on the timing of healthcare events, whereas the interviews and questionnaires captured current and retrospectively reported wellbeing and functioning. Together, these data enabled us to identify the life periods during which phenotypic differences between the genetic subgroups were most apparent.

Below, we provide detailed descriptions of the variables used to construct the heatmap in **Fig. 3** of the main manuscript. Interview and questionnaire items were grouped according to the life period and conceptually related items were combined into composite variables to reduce redundancy and capture broader dimensions that might not be adequately represented by individual items.

Composite scores were calculated as the sum of their component items. No phenotype imputation was performed. Because several composite variables had skewed distributions, group differences were evaluated using empirical Monte Carlo resampling where appropriate. For visualization, all variables were oriented so that higher values consistently represented less favorable outcomes and were subsequently transformed into rank-based deciles to place them on a common scale.

**Table SA2. Composite variables (Fig. 3 main manuscript)**

| **Early events** | | | |
|---|---|---|---|
| **Composite variable: Family situation** | | | |
| Variable | Range | Interpretation | Response rate |
| Separated parents | 0-1 | No / Yes | 96 % |
| Did your family have financial difficulties? | 0-1 | No / Yes | 86 % |
| Were your parents unemployed? | 0-1 | No / Yes | 91 % |
| Did any of your parents have a serious injury or illness? | 0-1 | No / Yes | 90 % |
| Did your mother and/or your father have alcohol problems? | 0-2 | No / One parent / Both parents | 91 % |
| Were there serious conflicts in your family? | 0-1 | No / Yes | 86 % |
| Did your mother and/or your father have a psychiatric illness? | 0-2 | No / One parent / Both parents | 82 % |
| **Composite variable: Performance in school** | | | |
| Self-rated school performance | 0-4 | 0=good, 2=average, 4=poor | 98 % |
| *Difficulties with: Mathematics | 0-1 | No / Yes | 97 % |
| *Difficulties with: Writing | 0-1 | No / Yes | 97% |
| *Difficulties with: Languages | 0-1 | No / Yes | 97% |
| *Difficulties with: Concentration | 0-1 | No / Yes | 97% |
| Need to repeat a grade | 0-1 | No / Yes | 97% |
| *A screening question asking whether difficulties had persisted for more than one year was used. Participants answering "No" did not receive the four follow-up questions and were here recorded as "0". | | | |

| Composite variable: Wellbeing in childhood | | | |
|---|---|---|---|
| Were you chronically ill in childhood? | 0-1 | No / Yes | 92 % |
| Were you bullied at school? | 0-1 | No / Yes | 94 % |
| Were you physically abused in childhood | 0-1 | No / Yes | 95 % |
| Were you sexually abused? | 0-1 | No / Yes | 93 % |
| Did you have behavior problems? | 0-1 | No / Yes | 50 % |
| **Adult and disease course events** | | | |
| **Composite variable: Attainment of adult social milestones** | | | |
| Have you been married? | 0-1 | No / Yes | 99 % |
| Do you have children? | 0-1 | No / Yes | 98 % |
| Do you have an occupation? | 0-1 | No / Yes | 96 % |

**Table SA3. Additional register-based and cross-sectional variables used in Fig. 3**

| Early events | |
|---|---|
| Variable | Definition |
| Age of first non-psychiatric hospitalization | Age at first hospital admission with a non-psychiatric primary diagnosis. |
| Age of first psychiatric hospitalization | Age at first hospital admission with a primary psychiatric diagnosis (i.e ICD 10 equivalent F.XX). |
| Avg psychiatric hospital burden (prior to psychotic diagnosis) | Average annual psychiatric hospitalization burden before the first register-recorded psychotic diagnosis. Details previously published (9). |
| **Adult / disease course events** | |
| Education (highest level) | 0=Basic/low, 1=secondary/middle, and 2=high education attainment. |
| Avg psychiatric hospital burden (post psychotic diagnosis) | Average annual psychiatric hospitalization burden after the first register-recorded psychotic diagnosis. Details previously published (9). |
| Age at first psychotic diagnosis | Age at first register-based diagnosis indicating psychosis. |
| Function at discharge | Mean Global Assessment Scale (GAS) score at discharge. |
| Clozapine use | No / Yes |
| Cognitive performance (PAL-errors) | Adjusted number of PAL errors after rank-based inverse normal transformation. |
| Supported housing | No / Yes |

**Comments:**

1) Throughout the study, several endpoints were defined using hospital admissions rather than all healthcare visits. This approach improved comparability across generations because only the Hospital

Discharge Register has provided nationwide coverage since 1969. Further details on register coverage are provided above (p. 2).

2) Because a psychotic disorder was required for inclusion in SUPER-Finland, age at psychotic illness onset was broadly defined as age at the first registry-recorded diagnosis indicating a psychotic illness. This should be interpreted as a register-based proxy for onset rather than the exact onset of symptoms. Previous analyses in SUPER-Finland support the validity of this approach (9).

ICD 8-10 definition of first sign of a psychotic disorder: ^F2[0-9]|^295[0-9][A-Z]|^29[7-8][0-9][A-Z]|^295[0-9][0-9]|^29[7-9][0-9][0-9]|^F30$|^F30[2-9]|^F31|^F3[2-3]3|^2960[A-Z]|^296[2-7][A-Z]|^296[1-9][0-9]|^F1007|^F1[0-9]0[3-4]|^F1[0-9][5-9]|^291[0-4][A-Z]|^292[1-9][A-Z]|^291[0-9][0-9]|^F0[4-5]|^F06[0-2]|F068|^F079|^F09|^293[0-9][A-C]|^29[2-3][0-9][0-9]|^294[0-2|4-9][0-9]|^F00|^F01|^F02|^F03|^290|^2941A|^29430|^2961E|^F531

**Table SA4. Variant carriers vs non-carriers (Fig. 3, main manuscript)**

| Variable | Statistical test | p-value | FDR adjusted p-value |
|---|---|---|---|
| **Early events** | | | |
| Childhood family circumstances | resampling | 4.18e-01 | 4.87e-01 |
| Age at first non-psychiatric hospitalization | resampling | 5.25e-03 | **1.21e-02** |
| Childhood wellbeing (self-rated) | resampling | 2.00e-05 | **1.40e-04** |
| School performance (self-rated) | resampling | 1.95e-03 | **5.46e-03** |
| Psychiatric hospitalization burden (prior to psychosis diagnosis) | resampling | 4.77e-02 | 6.07e-02 |
| Age at first psychiatric hospitalization | linear regression | 7.02e-05 | **3.28e-04** |
| **Adult / disease course events** | | | |
| Highest educational level | resampling | 6.06e-03 | **1.21e-02** |
| Age at first recorded psychotic diagnosis | linear regression | 4.30e-02 | 6.02e-02 |
| Attainment of adult social milestones | resampling | 2.60e-04 | **9.10e-04** |
| Psychiatric hospitalization burden (after psychosis diagnosis) | linear regression | 6.10e-01 | 6.57e-01 |
| Functioning at discharge | linear regression | 9.69e-01 | 9.69e-01 |
| Clozapine use | logistic regression | 3.83e-02 | 5.96e-02 |
| Cognitive performance (PAL errors) | linear regression | 1.20e-06 | **1.69e-05** |
| Supported housing | logistic regression | 1.11e-02 | **1.95e-02** |

Statistical significance was estimated using covariate-adjusted linear or logistic regression when the outcome distribution was suitable for regression-based inference. For outcomes with distributions not suitable for regression analysis, we used empirical Monte Carlo resampling. Groups of 163 eligible non-carriers were sampled without replacement 100,000 times, and the mean of each resampled group was compared with the observed mean among variant carriers. FDR-adjusted p-values were calculated across the 14 variables using the Benjamini–Hochberg procedure. All variant carriers had available values for the outcomes analyzed using Monte Carlo resampling. PAL performance, for which data were missing for some participants, was analyzed separately using linear regression.

**Direct effect size comparison between variant carriers and individuals with high SZ-PGS (≥90$^{th}$ percentile)**

The longitudinal analysis presented in **Fig. 3** of the main manuscript showed that variant carriers and individuals with high SZ-PGS had broadly similar overall phenotypic profiles. However, the timing of the observed differences appeared to vary: differences among variant carriers were most apparent before the first recorded psychotic diagnosis, whereas individuals with high SZ-PGS showed their largest differences later in the disease course. To compare these profiles directly, we performed a bootstrap-based analysis of standardized mean differences (Cohen's d) across the 14 variables included in Fig. 3. These variables represented different life periods and functional domains, including childhood wellbeing, the timing of clinical events, and later disease-course measures. Because several variables were composite measures or had skewed distributions, we used a nonparametric bootstrap approach. Results are displayed in Fig. S4.

For each variable, we calculated the difference between the observed mean among variant carriers and that among individuals in the high SZ-PGS group. Because the variables were measured on different scales, each mean difference was standardized using the pooled standard deviation of the two groups and expressed as Cohen's d. Uncertainty was estimated by resampling participants with replacement within each group 5000 times, and 95% confidence intervals were derived from the resulting bootstrap distributions. Positive values indicated less favorable outcomes among variant carriers because all outcomes had been oriented so that higher values represented less favorable outcomes. Two-sided p-values were estimated from the bootstrap distributions and adjusted across the 14 variables using the Benjamini–Hochberg FDR procedure.

The bootstrap comparison indicated that differences between variant carriers and individuals with high SZ-PGS were most apparent for childhood wellbeing, school performance, and age at first non-psychiatric hospital admission. These findings are consistent with the interpretation that damaging variants in SCHEMA-nominated SZ-risk genes are associated with more pronounced early-life differences than high SZ-PGS.

**Comparison between variant carriers and individuals with high SZ-PGS (Downsampling analysis)**

As displayed in Fig. 3 of the main manuscript, individuals with the highest SZ-PGS showed less favorable disease trajectories than reference non-carriers, primarily from the transition to psychotic illness onward. In the full analysis, high SZ-PGS was associated with more outcomes than variant carrier status. However, because the high SZ-PGS group was substantially larger, this broader pattern of statistically significant associations could reflect greater statistical power rather than larger phenotypic differences. We therefore downsampled the high SZ-PGS group to the same analysis sample size as the variant-carrier group (n = 163) to assess whether the high SZ-PGS associations remained detectable at a sample size comparable to that of the variant carriers. The results showed that only the association with age at first recorded psychotic diagnosis could be considered robust to downsampling (≥80% of iterations) and remained FDR-significant in 89.3% of the downsampled analyses. Age at first psychiatric hospitalization, clozapine use, psychiatric hospital burden (after first psychotic diagnosis) and cognitive performance remained moderately robust to downsampling and were significant in more than half of the downsampled analyses.

**Table SA5. High SZ-PGS group vs non-carriers (Fig. 3, main manuscript)**

| Variable | Full analysis | Downsampled analysis | |
|---|---|---|---|
| | FDR-adjusted p-value (high SZ-PGS group, n=958) | Proportion of iterations with FDR-adjusted p-value <0.05 | Median FDR-adjusted p-value (downsampled group, n=163) |
| Age at first psychotic diagnosis | 2.00e-05 | **0.893** | **7.00e-03** |
| Age at first psychiatric hospitalization | 2.00e-05 | **0.69** | **1.87e-02** |
| Clozapine use | 2.00e-05 | **0.687** | **2.04e-02** |
| Psychiatric hospital burden (after first psychotic diagnosis) | 2.00e-05 | **0.6** | **3.51e-02** |
| Cognitive performance (PAL-errors) | 2.00e-05 | **0.53** | **4.28e-02** |
| Attainment of adult social milestones | 2.00e-05 | 0.49 | 5.60e-02 |
| Highest educational level | 2.00e-05 | 0.37 | 9.06e-02 |
| Supported housing | 1.92e-04 | 0.27 | 1.43e-01 |
| Functioning at discharge | 7.78e-04 | 0.207 | 1.73e-01 |
| Psychiatric hospital burden (prior to psychotic diagnosis) | 1.34e-01 | 0.063 | 4.34e-01 |
| Childhood wellbeing (self-rated) | 1.34e-01 | 0.06 | 4.61e-01 |
| Childhood family circumstances | 2.66e-01 | 0.023 | 5.05e-01 |
| School performance (self-rated) | 5.63e-01 | 0.01 | 6.29e-01 |
| Age at first non-psychiatric hospitalization | 9.87e-01 | 0 | 8.23e-01 |

Statistical significance was assessed for all outcomes using one-sided empirical Monte Carlo resampling of the group mean. For the complete high SZ-PGS group (n=958), 100,000 reference samples were drawn from eligible non-carriers. For the downsampling analysis, 300 random subsets of 163 high SZ-PGS participants were selected, sampling without replacement within each iteration. For each outcome and subset, the observed mean was compared with the means of 3,000 reference samples drawn without replacement from eligible non-carriers. Each reference sample matched the number of high SZ-PGS participants with non-missing data for that outcome in the corresponding analysis.

Empirical p-values were calculated as the proportion of reference means at least as high as the observed mean (with one added to both the numerator and denominator). P-values were adjusted across the 14 outcomes using the Benjamini–Hochberg procedure for each downsampling iteration. For each outcome, we reported the median FDR-adjusted p-value across the 300 iterations and the proportion of iterations with an FDR-adjusted p-value <0.05. Three hundred iterations were used to maintain computational feasibility for this nested resampling procedure.

**Phenotypic endpoint definitions**

**Non-psychiatric causes of childhood hospital admissions.** Because hospital admissions with non-psychiatric primary diagnoses were much more common than admissions with psychiatric primary diagnoses, we examined whether their underlying causes differed across the genetic subgroups. Non-psychiatric primary diagnoses also encompassed a broader range of conditions than psychiatric diagnoses. We therefore grouped admissions into broad ICD-10 chapter-level diagnostic categories. Diagnoses recorded using ICD-8 and ICD-9 were harmonized to the corresponding ICD-10 categories. The analysis focused on the 10 most common non-psychiatric diagnostic categories up to 15 years of age. For each category, hospitalization burden was calculated by summing age-specific mean inpatient days, using the number of participants at risk for each one-year age interval as the denominator. The table below presents the ICD-10 categories and their corresponding ICD-8 and ICD-9 code mappings.

| ICD-10 top-level diagnosis category | ICD-10 codes | ICD-9 codes | ICD-8 codes |
|---|---|---|---|
| Some infectious diseases and parasitic diseases | A00-B99 | 001-139 | 000-136 |
| Tumors | C00-D48 | 140-239 | 140-239 |
| Diseases of the blood and blood-forming organs as well as certain disorders of the immune system | D50-D89 | 280-289 | 280-289 |
| Endocrine diseases, nutritional disorders and metabolic diseases | E00-E90 | 240-279 | 240-279 |
| Mental illness and syndrome as well as behavioral disorders | F00-F99 | 290-319 | 290-315 |
| Diseases of the nervous system | G00-G99 | 320-389 | 320-389 |
| Diseases of the eye and nearby organs | H00-H59 | NA | NA |
| Diseases of the ear and mastoid | H60-H95 | NA | NA |
| Diseases of the circulatory system | I00-I99 | 390-459 | 390-459 |
| Respiratory diseases | J00-J99 | 460-519 | 460-519 |
| Diseases of the digestive organs | K00-K93 | 520-579 | 520-577 |
| Diseases of the skin and subcutaneous tissue | L00-L99 | 680-709 | 680-709 |
| Diseases of the musculoskeletal system and connective tissue | M00-M99 | 710-739 | 710-738 |
| Diseases of the urinary and genital organs | N00-N99 | 580-629 | 580-629 |
| Pregnancy, childbirth and cot time | O00-O99 | 630-676 | 630-678 |
| Perinatal conditions | P00-P96 | 760-779 | 760-779 |
| Congenital malformations, deformities and chromosomal abnormalities | Q00-Q99 | 740-759 | 740-759 |
| Abnormal clinical and laboratory findings not elsewhere specified | R00-R99 | 780-799 | 780-796 |
| Injuries, poisonings and certain other consequences of external causes | S00-T98 | 800-999 | 800-999 |
| Codes for special purposes | U00-U99 | NA | NA |
| External causes of injuries, diseases and deaths | V01-Y98 | E | NA |
| Factors of importance for the state of health and for contacts with the health care system | Z00-Z99 | NA | NA |

**Register-recorded intellectual disability and neurodevelopmental disorders**

Register-recorded diagnoses indicating intellectual disability and/or neurodevelopmental disorders (ID/NDD) were defined using ICD codes corresponding to intellectual disability and autism spectrum disorders. In ICD-10, this corresponded to F70–F79 and F84.

The full regular expression used was: “^F7[0-9]|^F84|^31[7-9][0-9][A-Z]|^31[0-5][0-9][0-9]”.

The aim was to identify SUPER-Finland participants with register-recorded developmental diagnoses likely to be associated with cognitive impairment. This definition was intentionally narrow and was not intended to capture the full spectrum of neurodevelopmental conditions. The resulting endpoint was used only in sensitivity analyses assessing whether the association between variant carrier status and PAL test performance was driven by participants with register-recorded ID/NDD diagnoses.

**Definition of affective and non-affective diagnostic burden**

To examine whether variant carriers showed different affective and non-affective diagnostic profiles over time, we created two longitudinal, dimension-specific register datasets: one capturing non-affective psychotic diagnoses and one capturing affective diagnoses. These datasets were constructed from registry-recorded ICD codes in both inpatient and outpatient healthcare records. Outpatient records were included to increase power and to capture differences in longitudinal diagnostic profiles rather than only differences in the need for hospital care.

Non-affective psychotic diagnoses were defined as schizophrenia-spectrum and other non-affective psychotic disorders, excluding schizoaffective disorder. In ICD-10, this corresponded to F20–F24 and F26–F29. Corresponding ICD-8 and ICD-9 codes were also included. In the analysis script, non-affective psychotic diagnoses were identified using the following regular expression:

Non-affective psychotic diagnosis: ^F2[0-4]|^F2[6-9]|^295[0-6] |^295[8-9] |^29[7-8][0-9][A-Z] |^297[0-9][0-9] |^298[2-9][0-9] |^299[0-9][0-9].

Affective diagnoses were defined as all mood (affective) disorders and schizoaffective disorder. In ICD-10, this included F30–F39 and F25, irrespective of whether psychotic symptoms were recorded. Corresponding ICD-8 and ICD-9 codes were also included. Affective diagnoses were identified using the following regular expression:

Affective diagnoses: ^F3[0-9]|^2938D|^296[0-9][A-Z] |^296[0-9][0-9] |^298[0-1]0|^F25|^2957.

Because some inpatient episodes spanned several months, we did not treat each record as a single event occurring only on the admission date. Instead, inpatient episodes were expanded into calendar-month blocks covering every month from admission through discharge. For example, an inpatient episode beginning in January and ending in March contributed one record to January, one to February, and one to March. Outpatient records contributed to the calendar month in which the visit occurred.

After expansion into monthly blocks, records were deduplicated within each diagnostic dimension and calendar month. Thus, an individual could contribute at most once per month to each diagnostic dimension, even if multiple corresponding records occurred during that month. However, a long inpatient episode could contribute once for every calendar month it overlapped. Cumulative burden within each dimension was quantified as the total number of calendar months containing at least one corresponding diagnostic record. This approach captured the duration of long inpatient episodes while avoiding overcounting repeated healthcare contacts within a short period.

**References**

1. Sund R. Quality of the Finnish Hospital Discharge Register: a systematic review. Scand J Public Health. 2012;40(6):505-15.
2. Kurki MI, Karjalainen J, Palta P, Sipilä TP, Kristiansson K, Donner KM, et al. FinnGen provides genetic insights from a well-phenotyped isolated population. Nature. 2023;613(7944):508-18.
3. Trubetskoy V, Pardiñas AF, Qi T, Panagiotaropoulou G, Awasthi S, Bigdeli TB, et al. Mapping genomic loci implicates genes and synaptic biology in schizophrenia. Nature. 2022;604(7906):502-8.
4. Kaplanis J, Samocha KE, Wiel L, Zhang Z, Arvai KJ, Eberhardt RY, et al. Evidence for 28 genetic disorders discovered by combining healthcare and research data. Nature. 2020;586(7831):757-62.
5. PanelApp: Intellectual disability (Version 9.0). Accessed may 2025, URL: https://panelapp.genomicsengland.co.uk/panels/285/,.
6. Landrum MJ, Lee JM, Benson M, Brown GR, Chao C, Chitipiralla S, et al. ClinVar: improving access to variant interpretations and supporting evidence. Nucleic Acids Research. 2017;46(D1):D1062-D7.
7. ClinGen: https://search.clinicalgenome.org/kb/downloads#section_dosage, Accessed Sep 2024.
8. Karczewski KJ, Francioli LC, Tiao G, Cummings BB, Alföldi J, Wang Q, et al. The mutational constraint spectrum quantified from variation in 141,456 humans. Nature. 2020;581(7809):434-43.
9. Kämpe A, Suvisaari J, Lähteenvuo M, Singh T, Ahola-Olli A, Urpa L, et al. Genetic contribution to disease-course severity and progression in the SUPER-Finland study, a cohort of 10,403 individuals with psychotic disorders. Mol Psychiatry. 2024.
