## Supplementary Figures for "Ultra-rare coding variants in schizophrenia risk genes are associated with longitudinal phenotypic differences across psychotic disorders"

Fig. S1

### Comparison of psychotic diagnoses between the individuals with the lowest and highest SZ-PGS

a)

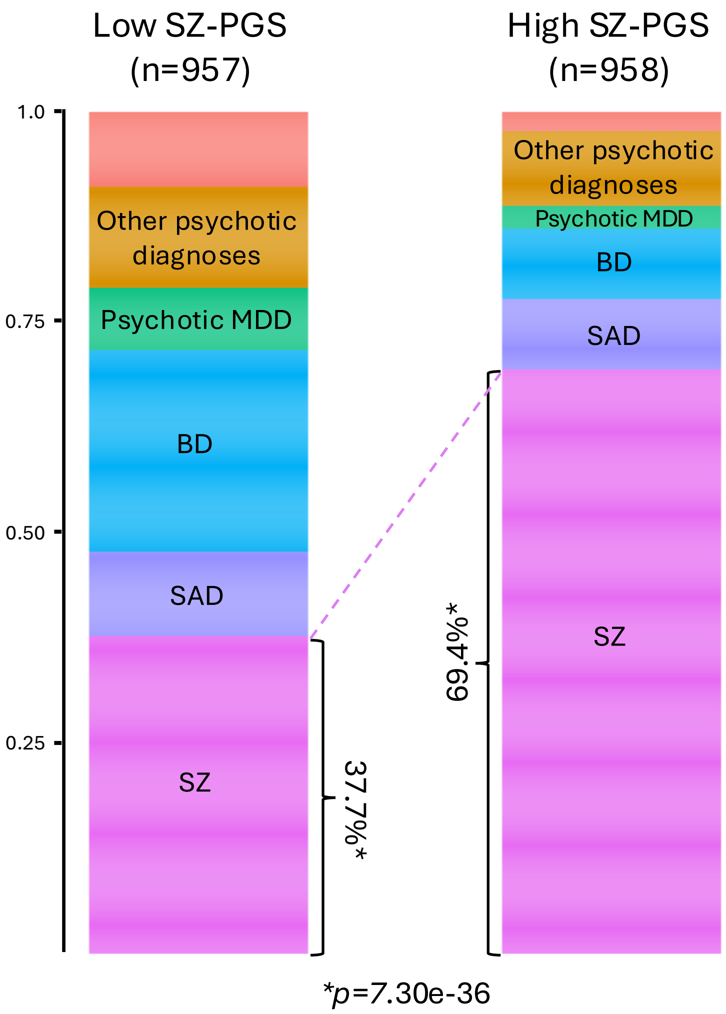


*Fig. S1a*. *Most severe psychotic diagnosis compared between individuals with the lowest (≤10th percentile) vs the highest (≥ 90th percentile) SZ-PGS of non-carriers in SUPER-Finland. Top bar displays the proportion without a psychotic diagnosis in the official healthcare registers.*

b)

| Parameter | Low SZ-PGS (≤10th percentile) | High SZ-PGS (≥90th percentile) | P-value |
|---|---|---|---|
| Year of Birth (mean) | 1971 | 1970 | 0.019 |
| Gender (male) | 45.7 % | 52.9 % | 1.46e-03 |
| **Psychotic diagnosis*** | | | |
| Schizophrenia | 37.7 % | 69.4 % | 7.30e-36 |
| Schizoaffective disorder | 19.3 % | 24.8 % | 2.44e-03 |
| Bipolar disorder | 33.3 % | 17.8 % | 1.42e-13 |
| Psychotic MDD | 21.3 % | 12.5 % | 5.63e-06 |
| Other psychotic Dx | 54.1 % | 67.3 % | 4.09e-10 |

*Psychotic diagnosis received at any timepoint. A single individual can have several different psychotic diagnoses.

*Fig. S1b*. *Demographic and diagnostic comparison between the individuals with the lowest SZ PGS (≤10th percentile) and the highest (≥90th percentile) SZ-PGS.*

Fig. S2

**Variant carriers: SCHEMA genes with a higher effect size showed lower SZ-PGS.**

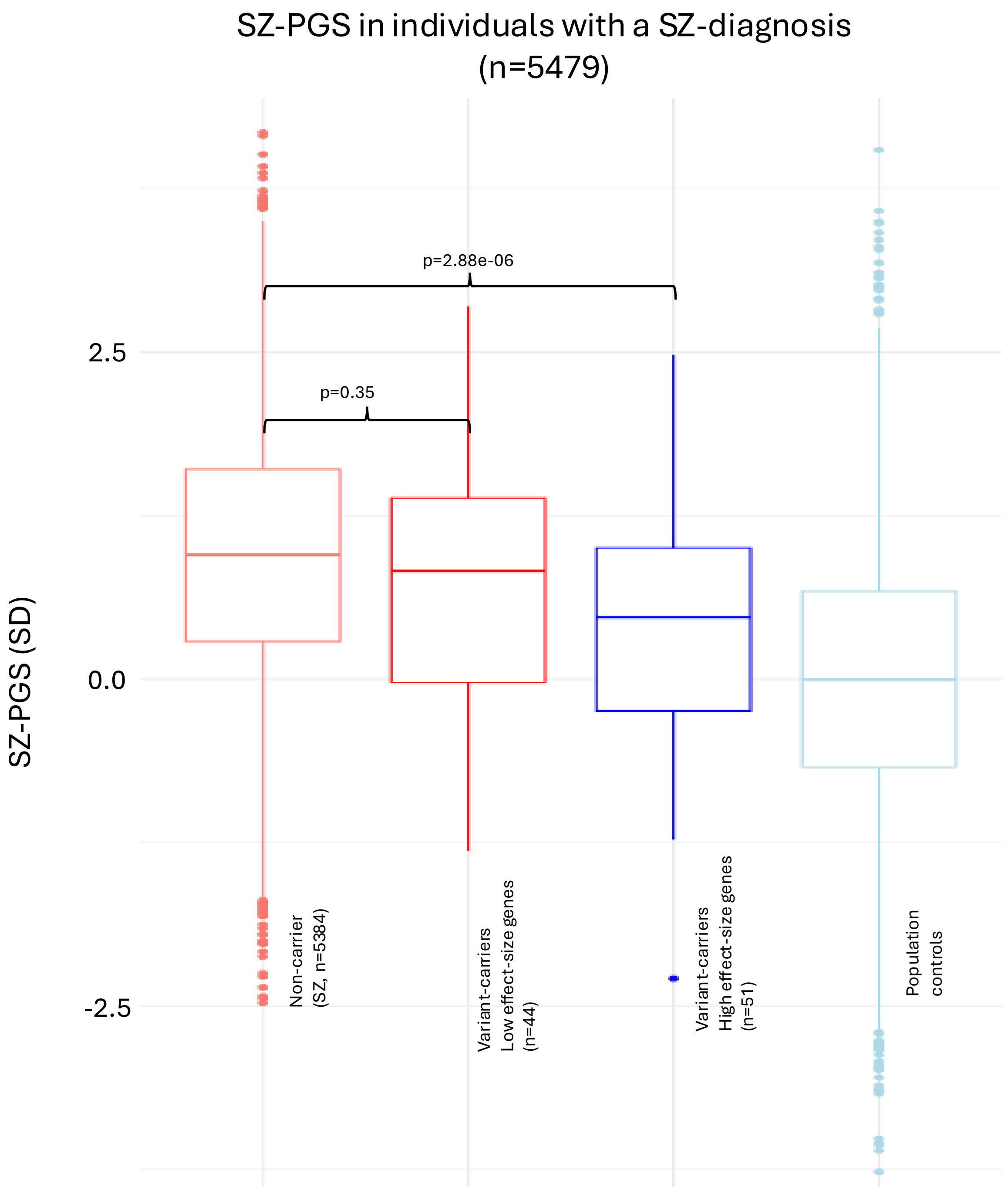


**Fig. S2.** The variant carriers had, on average, lower SZ-PGS than non-carriers. We observe that the variant carriers contributing the most to this signal had variants in genes with a higher effect size (top 50%) in the SCHEMA analysis.

*The analysis was restricted to individuals with a SZ diagnosis.*

Fig.S3

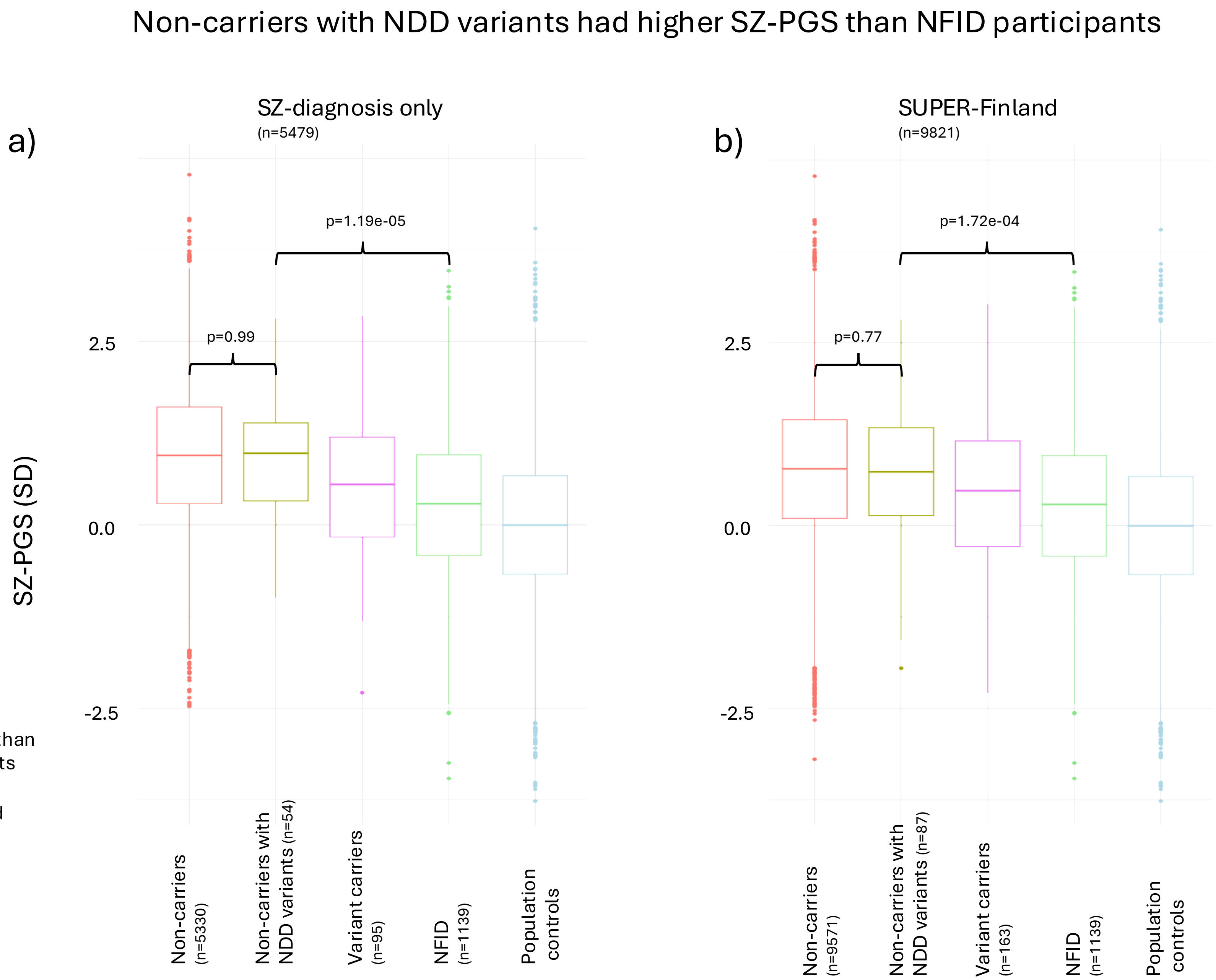


**Fig. S3.** Non-carriers with NDD variants had higher SZ-PGS than participants ascertained for ID in the NFID cohort. The results suggest that non-carriers with NDD variants, in contrast to SCHEMA variant carriers, require the same high background liability as the rest of the participants in SUPER-Finland to develop psychosis.

*Panel a) displays SUPER-Finland participants with a SZ diagnosis, while panel b) displays the entire SUPER-Finland cohort.*

Fig. S4

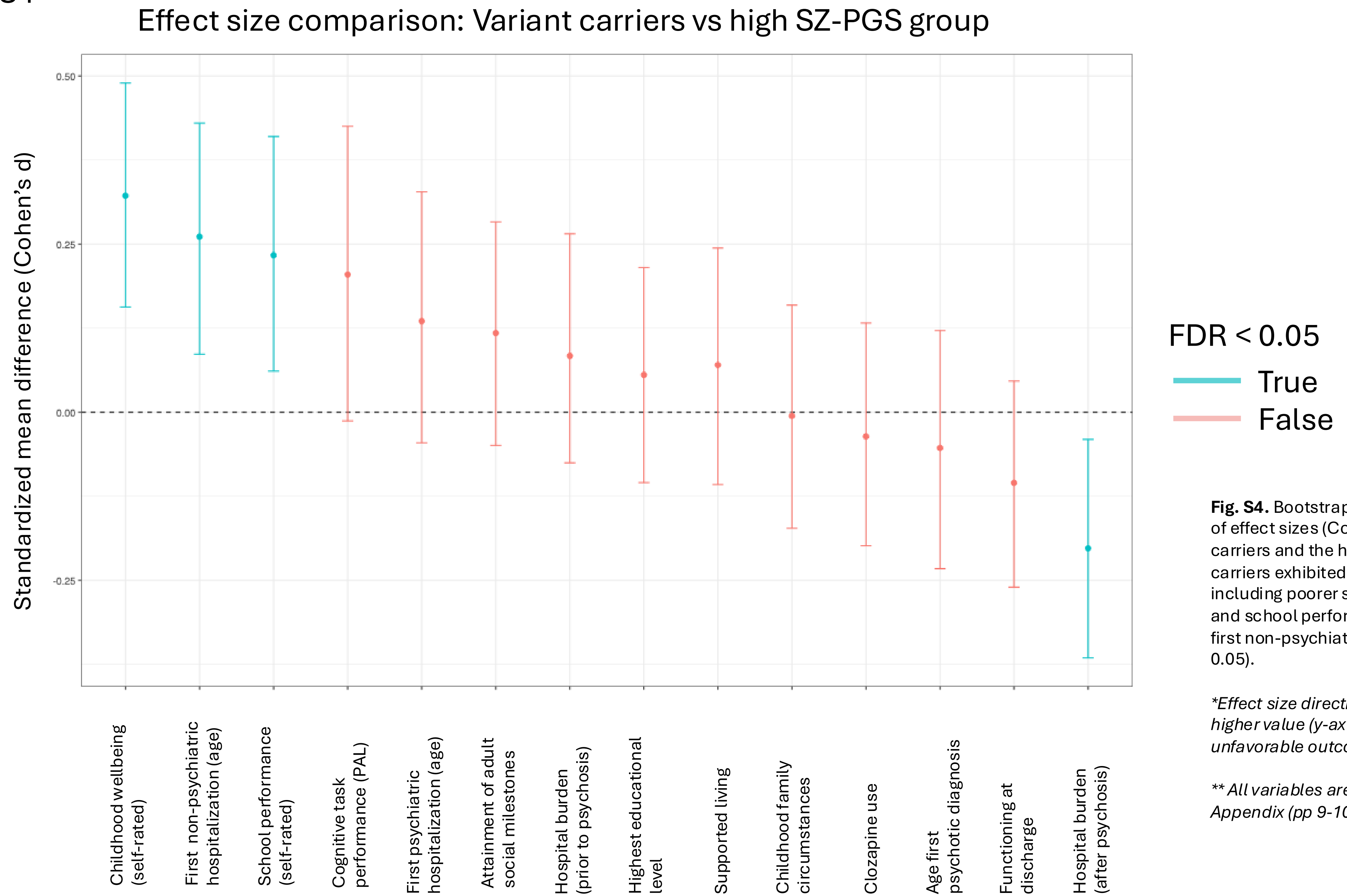


**Fig. S4.** Bootstrap-based group comparisons of effect sizes (Cohen's d) between variant carriers and the high SZ-PGS group. Variant carriers exhibited more adverse outcomes, including poorer self-rated childhood wellbeing and school performance, and an earlier age at first non-psychiatric hospital admission (FDR < 0.05).

**Effect size directions are aligned so that a higher value (y-axis) indicates a more unfavorable outcome for variant carriers.*

*** All variables are explained in detail in Supp. Appendix (pp 9-10).*

Fig. S5a

### Time to first psychiatric hospitalization

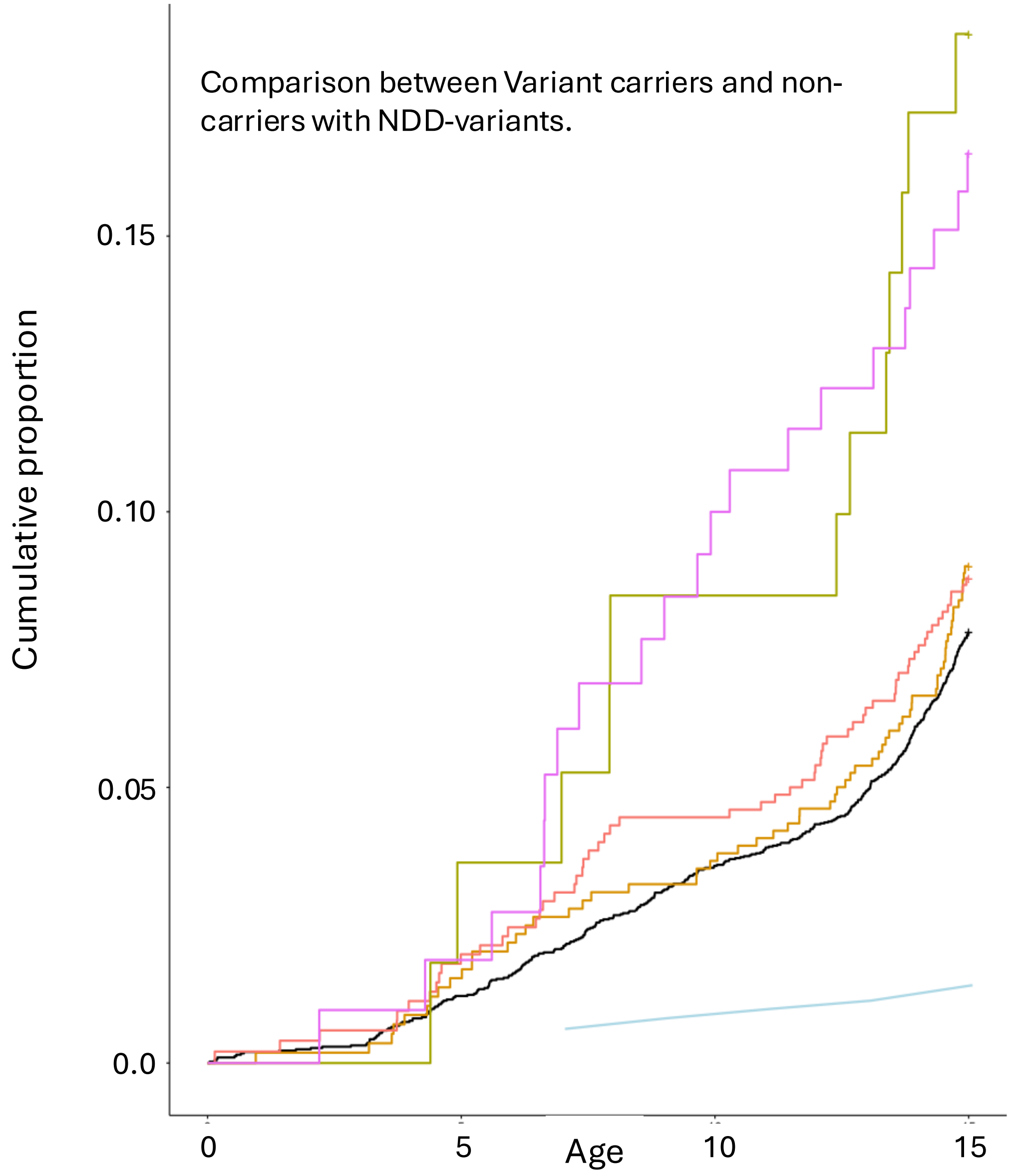


Subgroups (SUPER-Finland)

- Non-carriers (n=6363)
- Low SZ-PGS (10%, n=801)
- High SZ-PGS (90%, n=811)
- Non-carriers with NDD variants (n=69)
- Variant carriers (n=141)

**Numbers (n) reflect individuals eligible for the analysis, i.e., at risk after register start in 1969*. Individuals who had reached adulthood before register coverage began were not included.

| | |
|---|---|
| Non-carriers with NDD-variants (n=69) | HR=2.11 [1.18-3.76], p=0.01 |
| Variant carriers (n=141) | HR=2.30 [1.48-3.57], p=2.05e-04 |

*For reference*

FinnGen population (matched controls)

**Adjusted for sex, age, most severe psychotic diagnosis, PC1-10 and batch.*

**Fig.S5a.** Both variant carriers and non-carriers with NDD variants display earlier psychiatric hospital admissions compared to non-carriers. For the variant carriers the results remained unchanged also when ID-related diagnoses were excluded from the analysis (HR=2.28 [1.45-3.57], p=3.43e-04).

Fig. S5b

### Time to first non-psychiatric hospitalization

Subgroups (SUPER-Finland)

- Non-carriers (n=6363)
- Low SZ-PGS (10%, n=801)
- High SZ-PGS (90%, n=811)
- Non-carriers with NDD variants (n=69)
- Variant carriers (n=141)

**Numbers (n) reflect individuals eligible for the analysis, i.e., at risk after register start in 1969*. Individuals who had reached adulthood before register coverage began were not included.

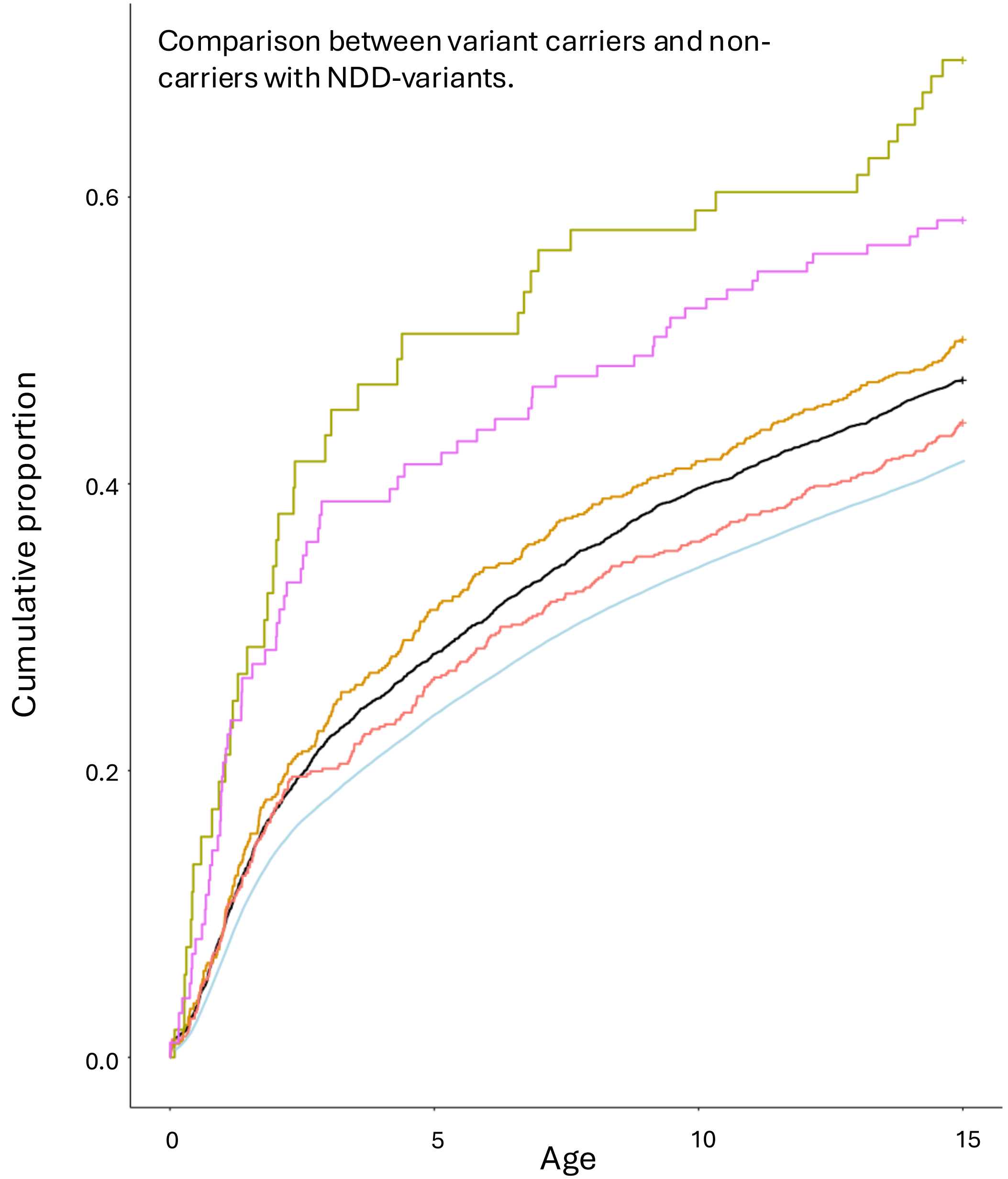


Non-carriers with NDD-variants (n= 69) HR=1.87 [1.37-2.53], p=6.69e-05

Variant carriers (n=141) HR=1.40 [1.10-1.79], p=6.88e-03

*For reference*

FinnGen population (matched controls)

**Adjusted for sex, age, most severe psychotic diagnosis, PC1-10 and batch.*

**Fig. S5b.** A similar pattern as for the variant carriers was seen for non-carriers with NDD-variants, displaying earlier hospitalizations due to non-psychiatric diagnoses.

Fig. S6

### In depth analysis of non-psychiatric hospitalizations

a) Most frequent non-psychiatric diagnostic categories (variant carriers, n=141 eligible)

*Only main diagnosis for hospital admission considered.*

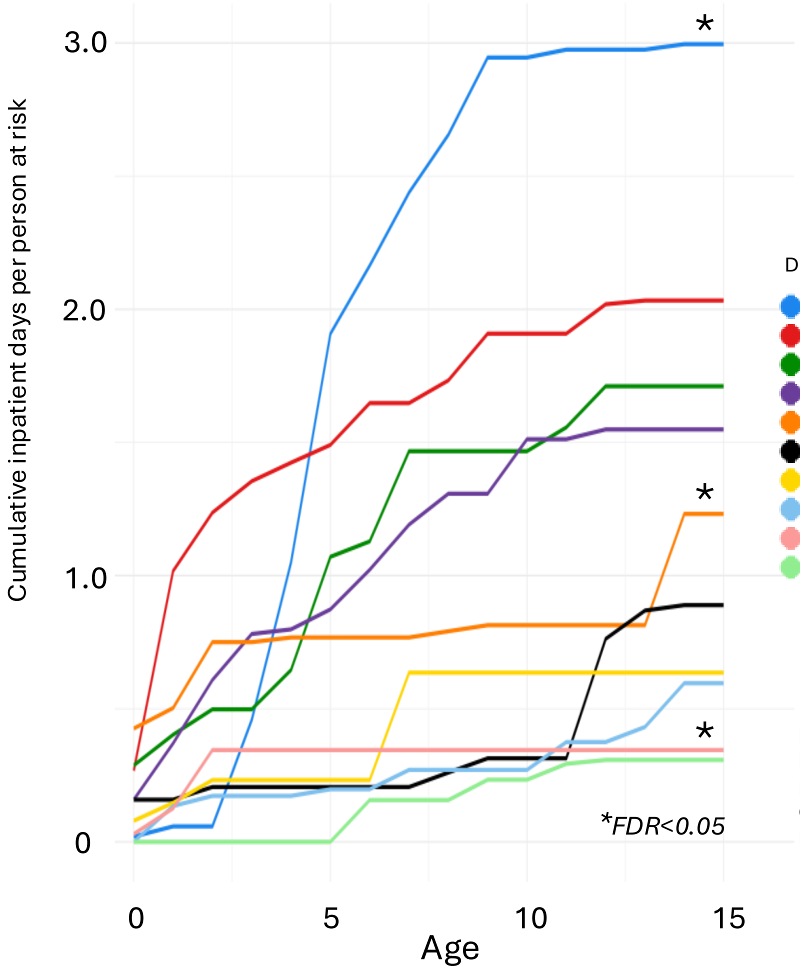


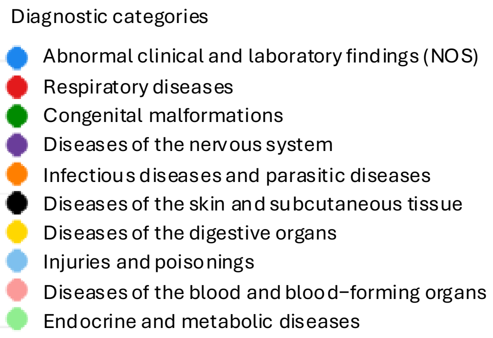


b) Monte Carlo simulation (resampling of non-carriers 10,000 times)

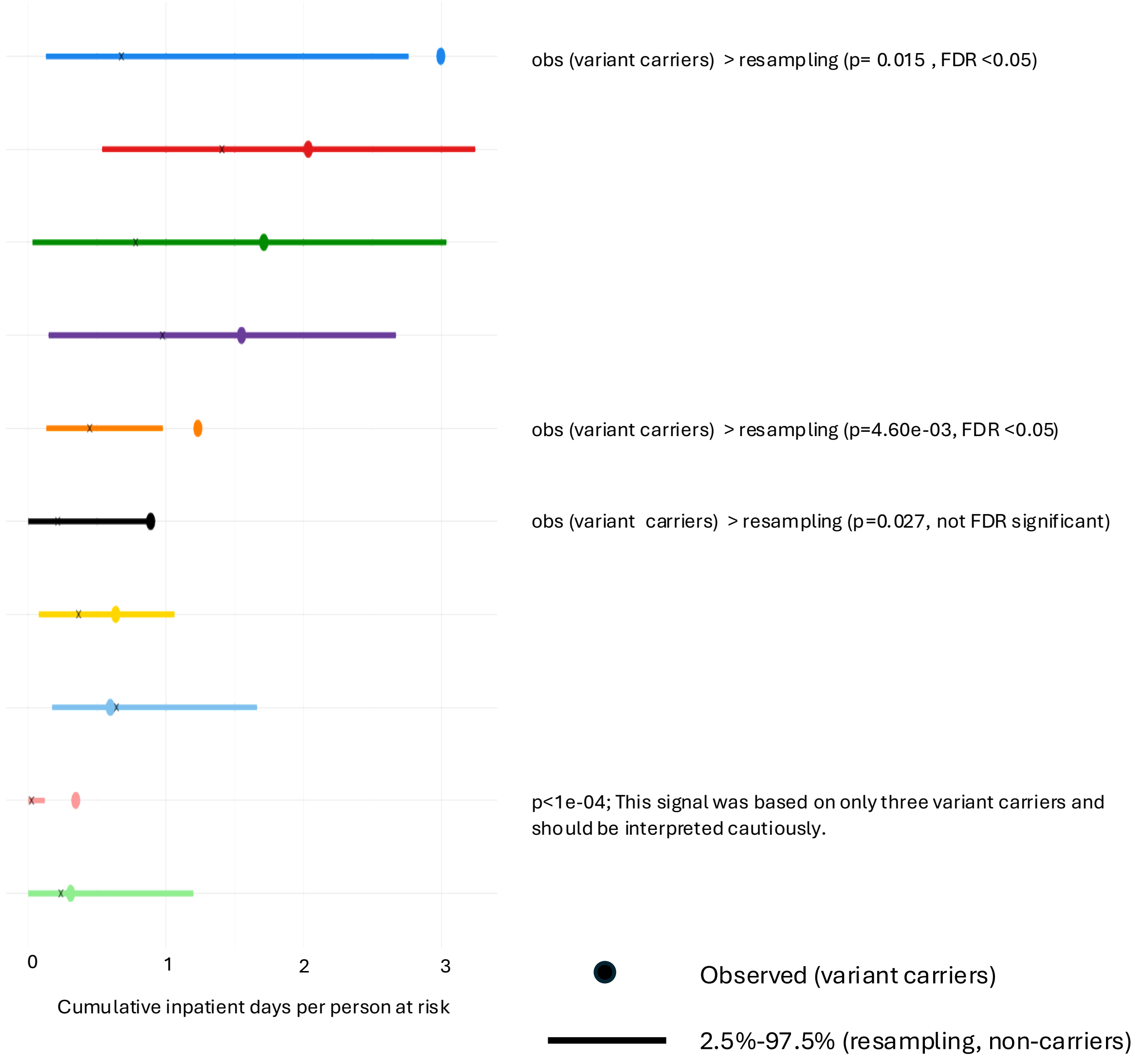


**Fig S6 a)** Primary reason for hospitalization for variant carriers (non-psychiatric diagnoses only). **b)** Comparison with non-carriers using resampling inference.

Fig. S7a-d

### Main diagnostic categories compared (non-psychiatric hospitalizations)

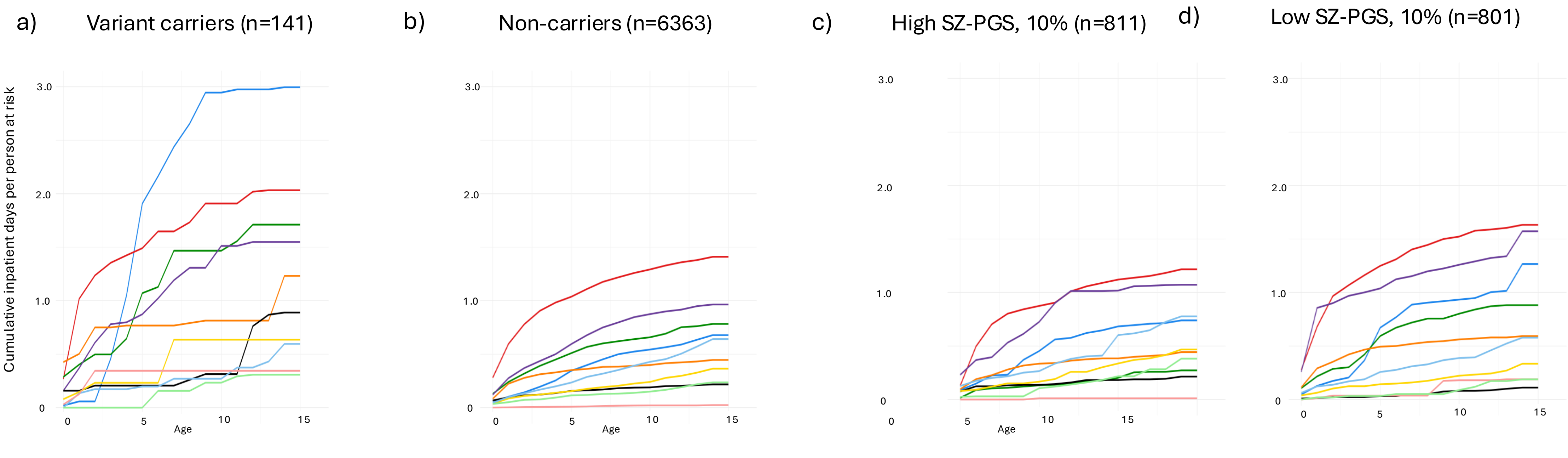


Diagnostic categories

- Abnormal clinical and laboratory findings (NOS)
- Respiratory diseases
- Congenital malformations
- Diseases of the nervous system
- Infectious diseases and parasitic diseases
- Diseases of the skin and subcutaneous tissue
- Diseases of the digestive organs
- Injuries and poisonings
- Diseases of the blood and blood–forming organs
- Endocrine and metabolic diseases

**Numbers (n) reflect individuals eligible for the analysis, i.e., at risk after register start in 1969.* Individuals who had reached adulthood before register coverage began were not included.

### Fig. S7e-g

#### Main diagnostic categories compared (non-psychiatric hospitalizations)

e) Non-carriers with NDD-variants (n=69)

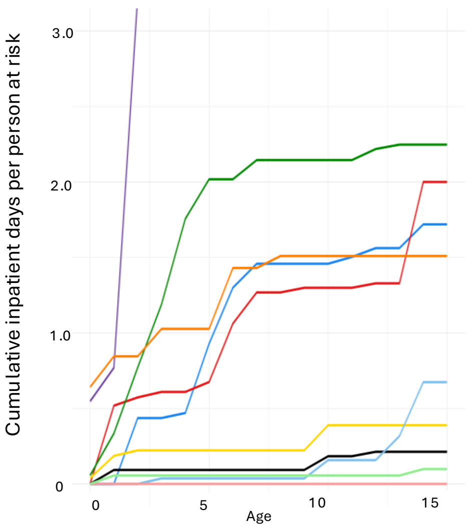


Scale change

f) Non-carriers with NDD-variants (n=69)

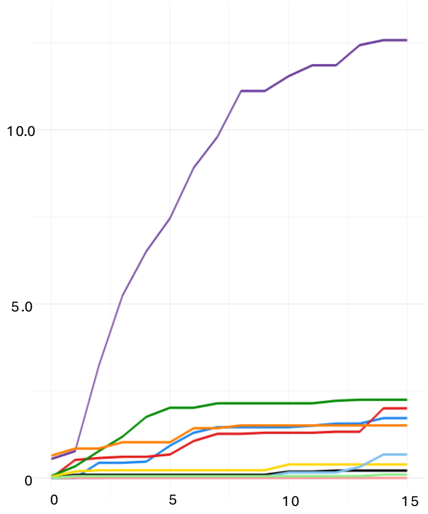


g) Monte Carlo simulation (Non-carriers with NDD-variants )

*Resampling of non-carriers 100,000 times*

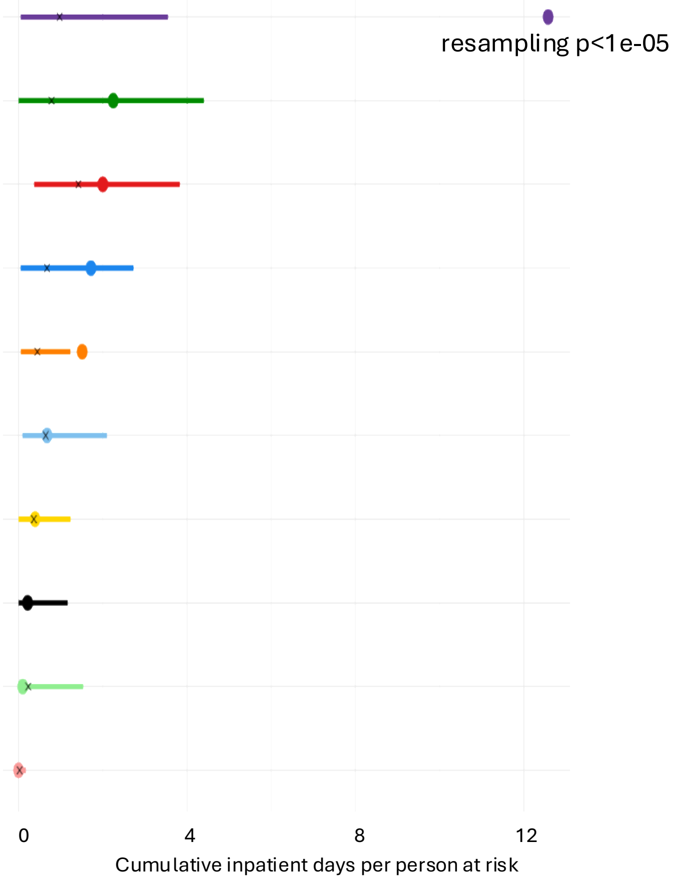


Diagnostic categories

- Abnormal clinical and laboratory findings (NOS)
- Respiratory diseases
- Congenital malformations
- Diseases of the nervous system
- Infectious diseases and parasitic diseases
- Diseases of the skin and subcutaneous tissue
- Diseases of the digestive organs
- Injuries and poisonings
- Diseases of the blood and blood−forming organs
- Endocrine and metabolic diseases

**Fig. S7.** Non-carriers with NDD variants displayed a clear enrichment of hospitalizations due to diseases of the nervous system. Panels e and f show the same results using different y-axis scales: panel e uses the scale applied in Fig. S7a–d, whereas panel f provides a zoomed-out view. Panel g compares non-carriers with NDD variants with other non-carriers. n denotes the number of eligible participants.

### Fig. S8

#### Association with PAL test performance (total errors) for different subgroups of variant carriers

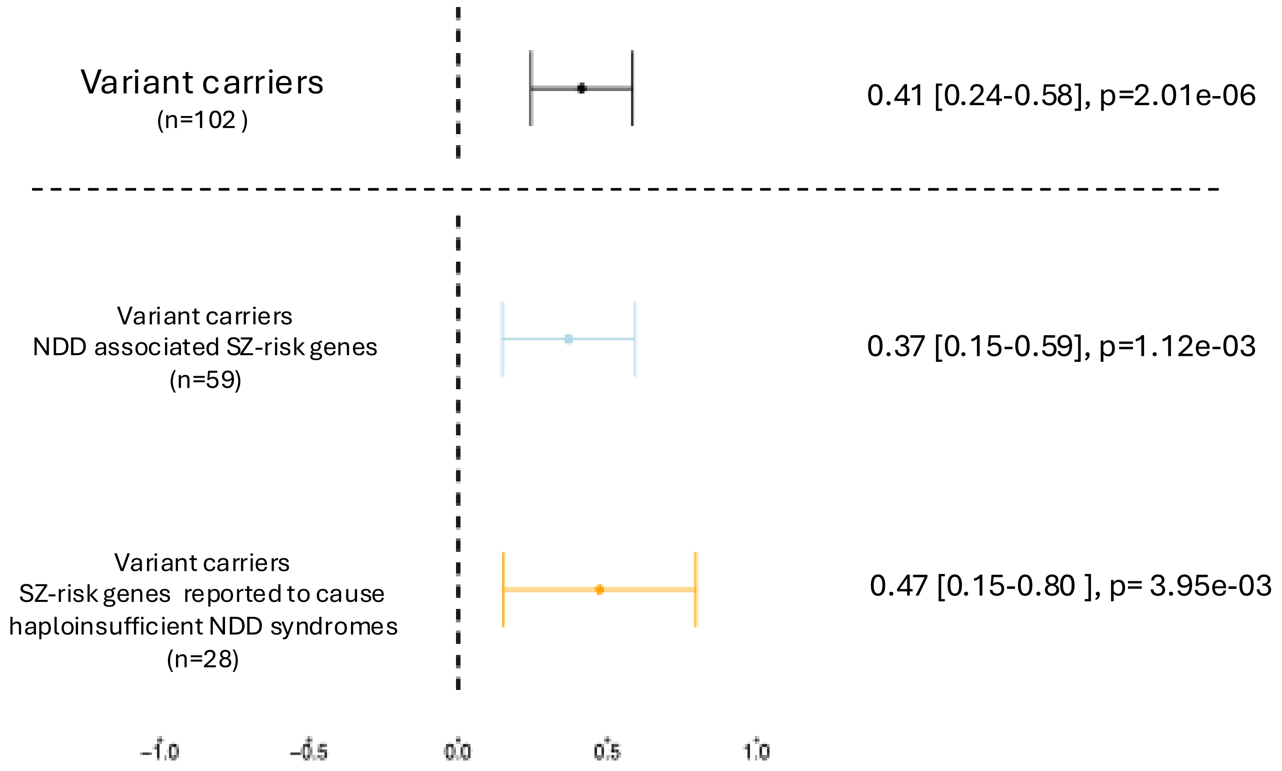


**Fig. S8.** Variant carriers whose affected SCHEMA-nominated genes have also been implicated in NDD syndromes performed similarly to other variant carriers on the PAL test. Variant carriers in SCHEMA-nominated genes previously reported to cause haploinsufficient NDD syndromes had a slightly higher point estimate, but this estimate did not differ significantly from that in other variant carriers ($p = 0.67$).

*n denotes the number of individuals eligible for the analysis, defined as those with a PAL test rated as "complete and reliable."*
